# Rare genetic coding variants associated with age-related episodic memory decline implicate distinct memory pathologies in the hippocampus

**DOI:** 10.1101/2024.05.21.24307692

**Authors:** Amanat Ali, Sofiya Milman, Erica F. Weiss, Tina Gao, Valerio Napolioni, Nir Barzilai, Zhengdong D. Zhang, Jhih-Rong Lin

## Abstract

**Background:** Approximately 40% of people aged 65 or older experience memory loss, particularly in episodic memory. Identifying the genetic basis of episodic memory decline is crucial for uncovering its underlying causes.

**Methods:** We investigated common and rare genetic variants associated with episodic memory decline in 742 (632 for rare variants) Ashkenazi Jewish individuals (mean age 75) from the LonGenity study. All-atom MD simulations were performed to uncover mechanistic insights underlying rare variants associated with episodic memory decline.

**Results:** In addition to the common polygenic risk of Alzheimer’s Disease (AD), we identified and replicated rare variant association in *ITSN1* and *CRHR2*. Structural analyses revealed distinct memory pathologies mediated by interfacial rare coding variants such as impaired receptor activation of corticotropin releasing hormone and dysregulated L-serine synthesis.

**Discussion:** Our study uncovers novel risk loci for episodic memory decline. The identified underlying mechanisms point toward heterogeneous memory pathologies mediated by rare coding variants.

## BACKGROUND

Cognitive aging is a natural process characterized by functional impairments across different cognitive domains including memory^1–3^. Episodic memory is a fundamental cognitive function that is particularly vulnerable to decline with aging^4,5^. Age-related episodic memory loss can manifest in various ways, including difficulties in learning and retaining new information. While some cognitive changes are considered manifestations of normal aging, progressive cognitive decline, especially in episodic memory, is a key clinical feature of Alzheimer’s disease (AD)^6,7^. Previous family and twin studies have demonstrated a moderate to high genetic influence on episodic memory in both cognitively healthy and cognitively impaired older adults^8,9^. However, further understanding of the genetic and molecular basis of episodic memory decline in the older population is needed to aid in the identification of novel drug targets to support cognitive reserve and, ultimately, to prevent and treat AD and related dementias.

In general, episodic memory reaches its peak in adulthood and may be subject to decline after the age of 60 years^5,10^. While memory function can be influenced by environmental and other factors that change with age^11^, the rate of memory loss appears to have a genetic component^12,13^. Previous genome-wide association studies (GWAS) of age-related cognitive decline ^14–16^, as well as episodic memory performance among older adults^17,18^, have identified AD risk loci with APOE being the major one; thus, prior findings strongly suggested a connection between AD pathology and episodic memory impairment in the general population. However, the discovery of other gene loci and their relationship to episodic memory decline has been relatively limited, despite experimental and clinical evidence that other factors, such as dysregulation of hormone and cerebrovascular supply^19,20,21,22^, may contribute to memory decline. Furthermore, the association of rare coding genetic variants with episodic memory decline remains largely unexplored. Uncovering these rare risk variants and understanding their underlying mechanisms may not only bridge existing research gaps but may also be of high scientific and clinical values, given the heterogeneity in pathological episodic memory decline in the population.

Employing a longitudinal study design, we tracked and analyzed the decline of episodic memory in 923 Ashkenazi Jewish participants from the LonGenity study, all age 60 years or older. We investigated both common and rare coding variants underlying individual differences in episodic memory decline using SNP-array and whole exome sequencing (WES) data, respectively. This study cohort is especially advantageous for identifying associations with causal rare variants at higher frequencies compared to the general population as it originates from a founder population of Ashkenazi Jews (AJs)^23^. Furthermore, AJs are relatively homogeneous in their education and socioeconomic status, which allows for better control of environmental factors that may impact memory decline^24^. Through genetic association analyses, we identified putative risk genes and pinpointed variants significantly associated with episodic memory decline. Subsequent post-GWAS and protein structural analyses were performed to gain insight into the biological mechanisms underpinning age-related episodic memory loss.

## METHODS

### Recruitment of study participants

LonGenity is a longitudinal study initiated in 2008 with the goal of identifying genotypes and phenotypes that protect against age-associated diseases. The LonGenity cohort is composed of older Ashkenazi Jewish individuals. Approximately half of LonGenity participants have a history of parental longevity, defined as having at least one parent who lived to age 95 years or older. Other prerequisites for eligibility include the absence of dementia at enrollment and age 60 or older (vast majority ≥ 65; mean age 75.4) (**Supplementary Fig. S1**). At annual visits, study participants are thoroughly characterized with evaluations that include medical history and neurocognitive assessments. Cognitive assessments were available for 1,112 (56.9% female) study individuals, but only 923 participants had at least two cognitive assessments. The Albert Einstein College of Medicine’s institutional review board (IRB) approved the LonGenity study.

### Episodic memory Assessment

Episodic memory was assessed at annual visits as part of a comprehensive cognitive battery. Standardized neuropsychological memory tasks (Weschler Memory Scale- R Logical Memory I and II^25^; Free and Cued Selective Reminding Test^26^ Free Recall and Total Recall, Recall of the RBANS figure)^27^ were administered and scored by the study research assistants under the supervision of the study neuropsychologist. As the tests were on different scales, we calculated Z scores standardized to baseline for the individual tests by subtracting the cohort mean score at baseline from the participant’s test score at each assessment and dividing by the standard deviation of the baseline scores. A composite memory score was generated by adding the 4 standardized memory scores.

### Measures of episodic memory decline

Since memory assessments at each study visit, as represented by standardized memory scores, were skewed, we transformed the standardized memory score phenotype using the log-based transformation (**Supplementary Fig. S2**):

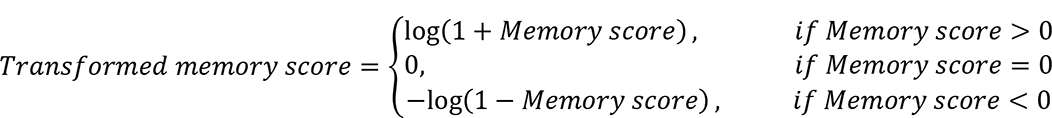

923 subjects had memory assessments for at least two waves. We measured the episodic memory decline of these 923 subjects by fitting a linear mixed model to the transformed memory scores of subjects at each wave. Baseline age (age at enrollment), sex, education years, and comorbidity score were included as predictors with fixed effects, while the number of years of follow-up after enrollment at each cognitive assessment was included as a predictor with mixed effects. The comorbidity score was defined as the sum of binary variables for the history of diagnoses of diabetes, high blood pressure, and stroke at the time of each cognitive assessment. The random effects of years of follow-up on the transformed memory score for each subject were collected as the measurement of the corresponding episodic memory decline (i.e., residual memory slope).

### SNP array genotyping

Among 923 study participants with memory measurements for at least two waves, only 791 had SNP genotype data. SNP genotyping was performed on two different genetic platforms. Regeneron and Center for Inherited Disease Research (CIDR) carried out genotyping using Illumina Global Screening Array-24 v.1.0 BeadChip with 642,824 markers and Illumina HumanOmniExpress-12v1 Array with 730,525 markers, respectively. A total of 629 study participants were genotyped on Illumina Global Screening Array-24 v.1.0 BeadChip and 162 individuals were genotyped on Illumina HumanOmniExpress-12v1 Array. Each SNP array genotyped dataset was ‘lifted over’ to human genome assembly GRCh38 separately. PLINK software (v.1.9) was used to perform quality control (QC) on each array-based genotyped data set separately. SNPs with minor allele frequency (MAF) < 1% were removed. SNP’s and sample’s missing rate was checked in two steps using >20% and >2% genotype calls threshold. SNPs and samples that passed these thresholds were retained. Study participants whose self-reported sex differed from that predicted by sex chromosome heterozygosity were excluded. SNPs were excluded if their genotype frequencies deviated from Hardy-Weinberg equilibrium by a χ2-test P < 1E–06. After that, samples were excluded if their heterozygosity varied from the mean by more than three standard deviations. A total of 150 CIDR genotyped and 602 Regeneron genotyped samples passed all QC steps.

### Exome sequencing and genotyping

The WES on samples from the LonGenity cohort was conducted by the Regeneron Genetics Center (RGC) using genotyping procedures as previously described^28^. After removing subjects with low sequencing coverage (less than 80% bases with coverage ≥ 20x), discordant sex, and call rate < 0.9, WES data was generated for subjects in the LonGenity cohort with at least two memory measurements. Genotypes called on human genome assembly GRCh38. We performed additional sample QC to remove subjects with a first-degree relationship (based on the identify-by-descent analysis) with another subject and ensured that all subjects had a call rate > 0.95 in the final study cohort that consisted of 632 subjects. For variant QC, the genotype of a SNP was not called if the read depth (DP) < 7 (DP < 10 for insertions/deletions (INDEL)). Variant sites were removed if the variant had a heterogeneous genotype for all subjects, but none of them had an alternative Allele Balance greater than a cutoff (≥ 15% for SNP, ≥ 20% for INDEL). We also removed variants from our analyses if their genotype frequencies deviated from Hardy–Weinberg equilibrium (χ^2^-test *P* < 1E–15). Only variants with missing rates < 0.01 in the study cohort were analyzed.

### Genotype imputation

Illumina Global Screening Array-24 v.1.0 (n = 602) and Illumina HumanOmniExpress- 12v1 Array (n = 150) genotyped datasets were independently imputed for genotype using the Michigan Imputation Server (Minimac4). For imputation, the Haplotype Reference Consortium (HRC, r1.1 2016) was used as the reference panel, the European (EUR) population for quality control and the Eagle v.2.3 for phasing. After imputation, variants that passed imputation reliability (R2 > 0.3) were retained for further analysis. Overall, 13,189k and 11,656K variants of genotyped datasets from the Illumina Global Screening Array-24 v.1.0 and Illumina HumanOmniExpress-12v1 Array, respectively, passed the imputation reliability (R2 > 0.3). A total of 11,084k variants were found to be common between the two datasets and were merged. A standard GWAS QC mentioned in SNP array genotyping section was again performed on the merged dataset. A total of 742 samples and 8,027k variants passed all QC steps and were used for downstream analysis. The imputation accuracy is high: in 66 subjects of which Whole Genome Sequencing (WGS) data is available, ∼98% of 1,100 randomly selected variants were accurately imputed (**Supplementary Fig. S3**).

### Common variant association analysis

A test of association between residual memory slope and autosomal variants was performed by using linear regression as implemented in PLINK v1.90. For this, only SNPs that passed MAF ≥ 1% were analyzed. We also filtered out duplicated variants and individuals with first-degree kinship. Baseline age, sex, years of education, the number of comorbidities, and the top 10 principal components from a principal component analysis (PCA) were included as covariates in the model. The number of comorbidities was defined as the sum of diagnoses history (0 to 3; 1 or 0 for each) for diabetes, hypertension, and stroke at or before the memory assessment. Risk loci were defined around genome-wide significant variants (<5 × 10^-8^); a region of ±250 kb was set around each variant. The PLINK clumping procedure was used to identify independent hits in each region. All variants were subjected to an iterative clumping procedure, beginning with the variant that has the lowest P value (known as the index variant). The index variant’s clump included variants with a P value (<5 × 10^-8^), located within 250 kb of the index variant and in linkage disequilibrium (LD) with the index variant (r2 above 0.1). After that, the clumping procedure was used to ensure that all of the variants were clumped.

### Rare variant association analysis

In our rare variant analyses, we only analyzed autosomal rare nonsynonymous variants. Rare variants are defined as variants of which alternative allele frequency < 1% in our study cohort. Coding variants were identified based on CADD annotation^29^ (v.1.6, ‘CodingTranscript’), in which nonsynonymous variants are those not annotated as ‘synonymous’ variants based on Ensembl Variant Effect Predictor (VEP). For single rare variant association analyses, we applied linear regression to test the association between individual variant genotypes and residual memory slope. Instead of testing the memory association of every rare coding variant, we ran the test on preselected rare nonsynonymous variants with the occurrence of at least five times in the study cohort and CADD scores ≥ 20. The reason for applying variant preselection is to focus on variants that are more likely to be functional and from which statistical tests can derive relatively reliable estimates while reducing the cost of multiple testing (**Supplementary note** and **Fig. S4**). For gene based rare variant association analyses, we ran SKAT-O^30^ to test association of putative functional rare nonsynonymous variants (CADD scores ≥ 20) on genes that met the following criteria: (1) genes that harbor at least two putative functional rare nonsynonymous variants to avoid redundant tests from single rare variant association analyses; and (2) genes with the summation of at least five occurrences of putative functional rare nonsynonymous variants observed in the study cohort from which statistical tests can provide relatively reliable estimates. Both single-variant and gene-based association tests included the following covariates: Baseline age, sex, years of education, the number of comorbidities, and top 10 principle components that account for population structure. The principal components were obtained using PLINK (v.1.9) based on common variants in the WES data. For both single and gene-based rare variant association tests, the false discovery rate (FDR) was used to account for multiple-test correction.

### The Alzheimer’s disease sequencing project (ADSP) study

We used WES data from the ADSP study to examine if carriers of rare coding variants had a higher chance of developing AD. The data consisted of 5,492 cases and 4,484 controls. We used Firth logistic regression to access the association between a rare variant and AD status; sex and top 10 principal components derived from common variants in the WES data were included in the regression as covariates.

### Replication study cohort

We prepared our replication study using people from the Religious Orders Study/Memory and Aging Project (ROSMAP)^31^ with both longitudinal episodic memory measurements (*n* = 3,771) and WGS data (*n* = 1,196)^32^ available. The episodic memory measurements at different visits for an individual were obtained from the Research Resource Sharing Hub (RADC) (https://www.radc.rush.edu/home.htm), and the WGS data was obtained from the AD Knowledge Portal (http://adknowledgeportal.org). We only included subjects without a diagnosis of AD and aged 65 or older at the first visit in the replication study to mimic the characteristics of our LonGenity study cohort (**Supplementary Fig. S1**). Furthermore, to achieve high-quality measurements of episodic memory decline and thus reliable replication results, we limited subjects to those having at least four annual episodic memory measurements, with an acceptable compromise of the sample size. Therefore, we recalculated residual memory slopes for subjects in our replication study – using the same method as in our discovery cohort – instead of using the existing measurements of episodic memory decline that have been generated from previous studies^14^. For the WGS data, only variants passing all filtering criteria were considered. The top 10 principal components accounting for the subpopulation structure were calculated based on common variants in the chromosome 22.

### Protein modeling

The top rare coding missense variants associated with episodic memory decline at the variant and gene levels were further evaluated for their impact on protein structure and function. A set of protein engineering approaches was employed to obtain molecular insights into how rare coding missense variants affect protein structure and function. First, homology modeling was performed to determine the three-dimensional (3D) structure of proteins (wild types and mutants) whose human 3D structure was not available in the protein database (PDB). Models were produced using Schrödinger Prime 2022-2 (Prime, Schrödinger, LLC, New York, NY, 2022), or AlphaFold^33^, where available. Loops refinement and structure validation of all 3D protein structures were carried out in Schrödinger Prime 2022-2 (Prime, Schrödinger, LLC, New York, NY, 2022). Schrödinger’s protein preparation wizard was used to exclude unnecessary water molecules and it also relaxed multimeric complexes, allocated bond orders accurately, produced disulfide bonds, corrected the orientation of misoriented groups, and adjusted ionization states. Standard protonation states at pH 7 were applied, and hydrogen atoms were added to the protein structures. Then, to produce geometrically stable structures, the preprocessed structures were optimized and minimized.

Second, the residue scanning tool in Schrödinger Prime 2022-2 (Prime, Schrödinger, LLC, New York, NY, 2022) was used in an implicit solvent model to determine the impact of mutations on protein stability. Third, the functional nature of variants was ascertained, particularly those that were directly involved in the disruption or formation of new critical intermolecular interactions identified via protein engineering approaches. Fourth, to confirm if a residue was present at an interface essential for protein-protein interaction (PPI), it was checked in computationally predicted interface residues of experimentally determined binary PPIs from Interactome INSIDER (v.2018.3, http://interactomeinsider.yulab.org). If crystal structures of proteins with known interacting partners were available in PDB, either as homodimers or heterodimers, they were used for further analysis. For others, protein-protein docking was performed using Schrödinger Protein-Protein Docking Suite 2022-2 (Schrödinger, LLC, New York, NY, 2022) to obtain the structure of a complex. For PPI crystal structures and generated docked complexes, any residue that was at the surface of a protein and whose distance to the interacting partner interfacial residue(s) was <5 Å was regarded as being at the interface.

To gain mechanistic insights into interfacial variants at an atomic level, all-atom molecular dynamics (MD) simulations were performed using Desmond^34^. For this, the 3D coordinates of CRHR2 bound to G proteins and PSPH were retrieved from PDB (PDB ID: 6PB1 and 1L8L), and structures were prepared using Schrödinger’s protein preparation wizard as described above. To obtain the phosphoserine bound complex of PSPH, phosphoserine was first docked to the well defined active site of PSPH using Schrödinger Glide’s Standard Precision (SP) method with default parameters (Schrödinger Glide, LLC, New York, NY, 2022). The best-docked pose of phosphoserine with the lowest Glide Score (GScore) value was used for molecular dynamic (MD) simulations and further analysis. Complex structure of PSPH was placed in an orthorhombic box of size 100 Å × 100 Å × 120 Å. Since CRHR2 is a GPCR, therefore, the bound structure of CRHR2 was embedded into a pre-equilibrated DPPC membrane in an orthorhombic box, with a buffer distance of 10 Å. Both systems were solvated with single point charge (SPC) water molecules using the Desmond System Builder (Schrödinger, LLC, New York, NY). The prepared simulation systems were neutralized with counter ions as well as 0.15 M salt concentration of NaCl was maintained.

All simulations were performed using Desmond. All calculations were carried out using an OPLS forcefield. Each prepared system was subjected to the default eight-stage relaxation protocol of Desmond. After relaxation, production simulation runs were performed for 500 ns in triplicates using different seed velocities. The pressure at 1 atm and temperature at 300 K, were maintained using isotropic Martyna–Tobias–Klein barostat and the Nose– Hoover thermostat ^35,36^, respectively, during the simulations. The smooth particle mesh Ewald (PME)^37^ method was used for long-range columbic interactions whereas short-range cutoff was set as 9.0 Å. MM-GBSA method was employed to estimate the free energy of binding of the G protein subunit to CRHR2 and phosphoserine ligand to PSPH using frames extracted from MD simulation trajectories. Every 100 ns, frames from each simulated system were extracted, and MM-GBSA based binding free energy was calculated using Schrödinger Prime employing the VSGB 2.0 solvation model^38^. Simulation data was analyzed using packaged and in-house scripts.

## RESULTS

### Memory performance in the study cohort

The study cohort of episodic memory decline included 923 individuals, each having a minimum of 2 memory assessments. On average, each participant completed 5.5 cognitive assessments over 6.2 years (**Supplementary Fig. S5**). Linear mixed regression model analysis of 5,034 memory assessments revealed a negative correlation between years of follow-up and memory scores (*P* = 3.41E-08), indicating that memory scores at later follow-up times tended to be lower. Older baseline age (age at enrollment) was also associated with lower memory scores (*P* < 2E-16). Females, on average, had higher memory scores than males (*P* = 5.86E-06). As expected, education years positively correlated with memory scores (*P* = 1.22E-10). There was no significant association between memory scores and the number of comorbidities in our cohort (*P* = 0.126). The measurement of episodic memory decline (residual memory slope) for each subject was derived as the random effect of years of follow-up in a linear mixed-effect regression model (see **Methods**) (**Fig. 1a** and **Supplementary Fig. S6.**). The residual memory slope exhibited a significant negative correlation with baseline age (*P* = 1.66E-06) (**Fig. 1b**), suggesting that individuals who enrolled at a later age were more likely to experience episodic memory decline in subsequent years. Conversely, we did not observe a significant correlation between the residual memory slope and sex (*P* = 0.409) or education years (*P* = 0.43) (**Fig. 1c** and **1d**). While participants varied in the number of cognitive assessments, the effect of practice/learning that results from exposure to the same tests multiple times was accounted for by adjusting for follow-up time, which was highly correlated with the number of cognitive assessments (correlation coefficient = 0.96) (**Supplementary Fig. S7**). Furthermore, we confirmed that the residual memory slope was not correlated with the number of assessments completed by each participant (*P* = 0.826), indicating that the learning effect was accounted for in the linear mixed model. Approximately half of our participants were offspring of parents with exceptional longevity (OPEL), and the OPEL exhibited a borderline positive association with the residual memory slope (*P* = 0.062) (**Supplementary Fig. S8**), suggesting that familial longevity may confer genetic protection against episodic memory decline. The derived residual memory slope from fitting linear mixed-effect regression in 923 individuals was used as phenotypes of subjects in the common and rare variant association analyses (*n* = 742 and 632, respectively) (**Supplementary Fig. S9**). The slope derived from the entire cohort was very similar to the slope derived from the cohort subsets used for common or rare variant analyses (correlation coefficient > 0.99).

**Figure 1.**
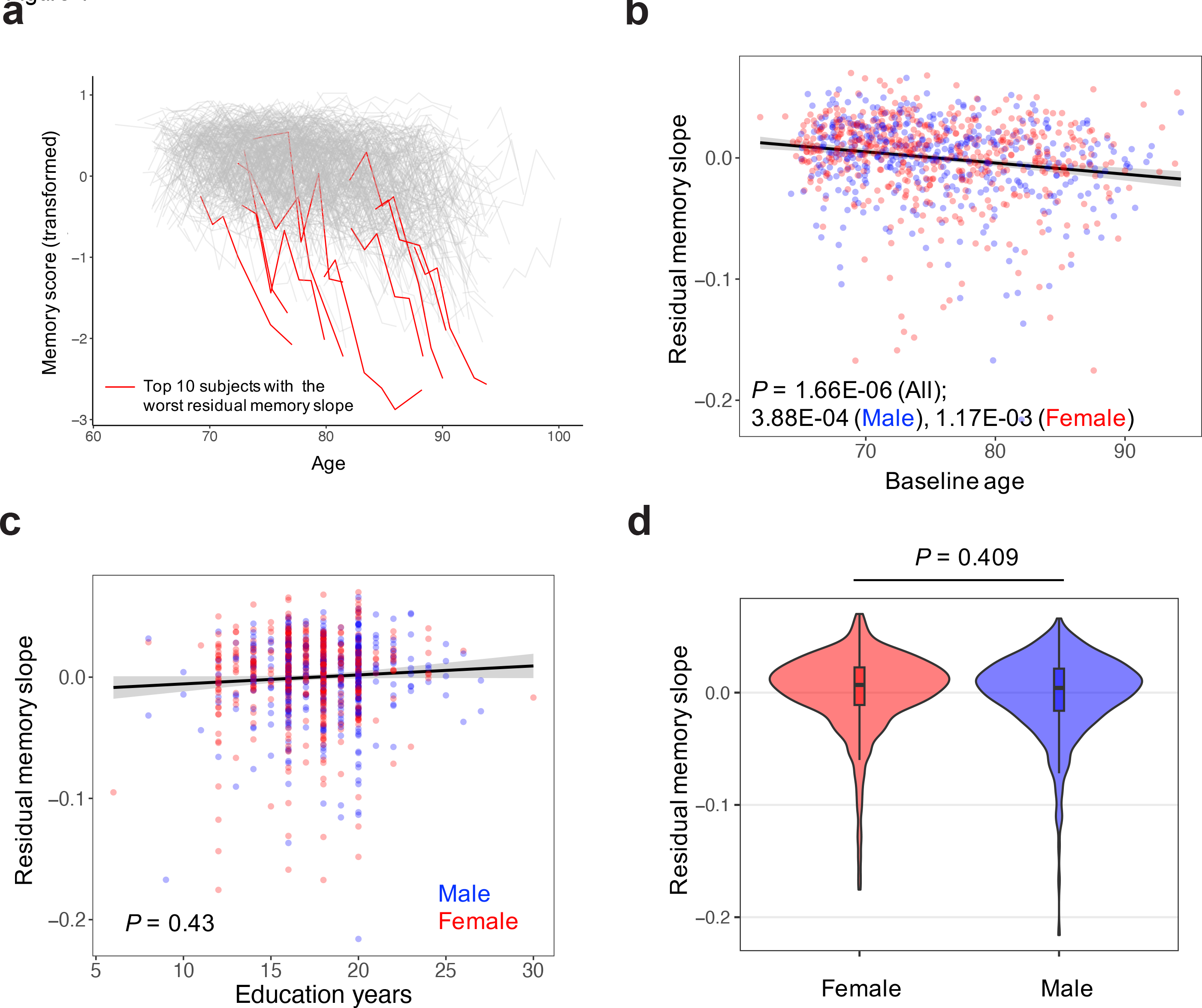
**Distribution of memory measures**. **a.** Memory trajectories of 923 subjects and 10 subjects with the worst residual memory slope (red) derived from the linear mixed model. **b-d.** The correlation between the residual memory slope and baseline age (b), education years (c), and sex (d).

### Common variant association analysis

The common variant study cohort consisted of 742 subjects from the LonGenity cohort, for whom both SNP array and residual memory slope data were available (**Supplementary Table S1**). We conducted GWAS on 8,027k common variants (MAF ≥ 1%) that passed QC after imputation (see **Methods**) and confirmed no inflation of test statistics (the genomic inflation factor λ = 1.01) (**Supplementary Fig. S10**). The association analysis identified three LD-independent genome-wide significant SNPs (*P* < 5E-08): rs548640610, rs35990795, and rs541421523 (**Fig. 2a** and **Table 1**). These three SNPs were not tag SNPs but had the strongest association among imputed SNPs in their genomic regions. Since none of these SNPs were located at coding regions, we examined the epigenetic information for these positions (**Fig. 2b-g**) using publicly available single cell Chip-seq and ATAC-seq data from different brain regions^39^ in order to ascertain the biological relevance of these putative causal SNPs. rs548640610 resides within the neuron and astrocyte-specific regulatory elements (**Fig. 2e**) and is located in an intron of *CHRM3*, which is expressed in the cortex region of the brain. rs548640610 was predicted to occur in a region with unusually strong enrichment for the binding of transcriptional coactivators (super-enhancers). Approximately 19% of AD-associated SNPs have been shown to occur in genome regions encompassed by brain tissue super-enhancers^40^. rs541421523 resides in the neuron, oligodendrocyte, and astrocyte-specific regulatory elements (**Fig. 2f**). rs35990795 resides in the microglia, oligodendrocytes, and astrocytes-specific regulatory elements (**Fig. 2g**) and is located in an intron of *CLIC6*. Interestingly, rs35990795 is eQTL for *CLIC6* in two brain regions based on the GTEx database^41^ (putamen, *P* = 7.0E-08; cerebellar hemisphere, *P* = 4.0E-05).

**Figure 2.**
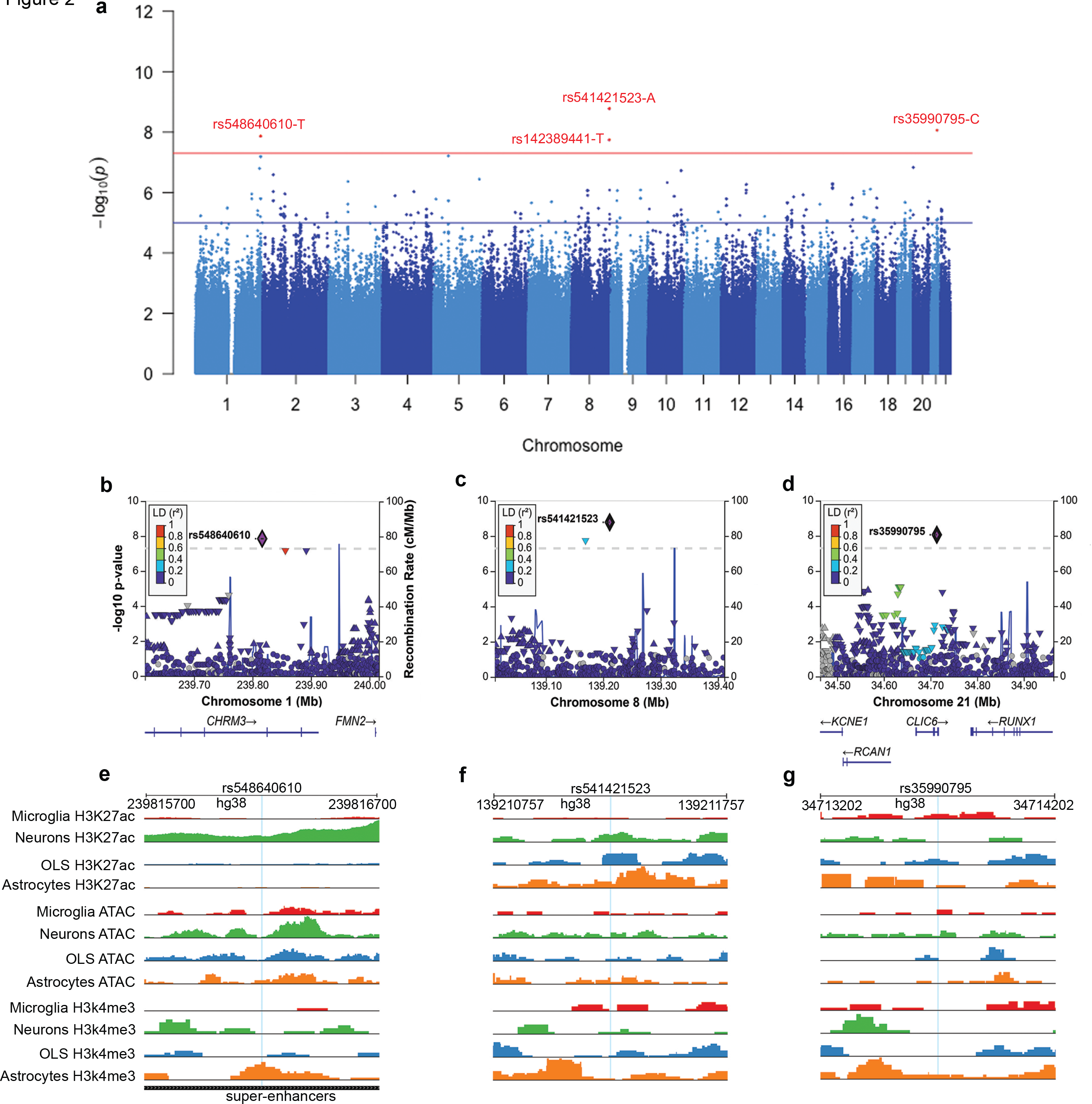
Common genetic variants associated with episodic memory decline. a. Manhattan plot demonstrating GWAS results for episodic memory decline. The red and blue lines represent thresholds for genome-wide significance (p < 5 × 10^-8^) and suggestive significance (P < 1 × 10^-5^), respectively. **b-d**. Detailed view of the GWAS SNPs demonstrating associations +/− 400 kb from lead SNPs (labeled). The X-axis represents the variant position on the chromosome and nearby gene positions. The right y-axis indicates the GWAS p-value, and the left y-axis indicates the rate of recombination. Each plot point indicates a SNP in the dataset color-coded by (r2) value using the EUR population from the 1000G LD panel. LD plots were generated using the Locus Zoom plot (locuszoom.org). **e-g**. Cell-type specific regulatory architecture of the following GWAS significant SNPs: rs548640610 (e), rs541421523 (f), and rs35990795 (g). A 1,000bp window flanking the SNPs is shown along with the genome tracks illustrating the aggregate accessibility of scChip-seq and scATAC-seq clusters at the locus. A blue line in Figure e-g indicates the location of GWAS significant SNPs.

### Analysis of AD common polygenetic risk

Since episodic memory loss is a prominent feature of AD, we hypothesized that some participants experienced episodic memory decline due to AD pathology. To test this hypothesis, we investigated whether the genetic risk for AD contributed to episodic memory decline in our cohort. First, we examined the APOE4 risk allele – the common risk variant with the strongest effect size in AD. However, we did not observe a significant association with the residual memory slope (*P* = 0.09), even though APOE4 carriers tended to exhibit a more negative residual memory slope. Next, we assessed in our cohort the common polygenic risk of AD, characterized by AD Polygenic Risk Score (PRS) based on the summary statistics of AD GWAS^42^. Using the default setting of LD-clumping in PRSice-2, we identified a small but significant component of variance for residual memory slope explained by AD PRS (*R*^2^ = 1.1%, adjusted *P* = 0.023) (**Supplementary Fig. S11a**). Given that the summary statistics of AD were derived from subjects predominantly of European ancestry, and the subjects in the target cohort of the PRSice-2 analysis were AJs, the standard LD-clumping procedures might fail to select optimal representative SNPs in a genomic region accounting for AD risk due to the discrepancy in LD structures between the two populations. This discrepancy might result in power loss in PRS analyses. Indeed, when using extremely parsimonious clumping parameters to select top SNPs in genomic regions without considering LD structures (clump-*kb* = 500*kb*, clump-*r*2 = 0), the result clearly showed that AD PRS contributed to episodic memory decline in the LonGenity discovery cohort (*R*^2^ = 2.6%, adjusted *P* = 2.0E-04) (**Supplementary Fig. S11b**).

### Rare variant association analysis

The rare variant study cohort consisted of 632 subjects from the LonGenity cohort, for whom both WES and residual memory slope data were available. (**Supplementary Table S1**). We identified a total of 116,438 rare coding variants (112,972 SNPs and 3,466 INDELs) with alternative allele frequency (AAF) < 1% across 17,208 genes in the study cohort. Among these, 40,653 were synonymous, and 75,785 were non-synonymous variants, which included 67,592 missense, 2,977 loss-of-function (stop gained and frameshift), and other variants with multiple annotations. We first used linear regression analysis to assess the association of these rare coding variants with residual memory slope. Next, we focused on 8,562 rare coding variants that had at least 5 occurrences in the cohort and had a CADD score ≥ 20, among which the derived significant associations were more reliable (**Supplement note** and **Fig. S4**). We confirmed that there was no systematic inflation (the genomic inflation factor λ = 0.98) (**Supplementary Fig. S12**) among those 8,562 rare coding variants and identified 18 putative risk rare coding variants (FDR < 0.05) associated with residual memory slope (**Fig. 3a** and **Supplementary Table S2**). All those putative risk rare coding variants increased the risk of episodic memory decline. For example, five out of six carriers of the top putative risk rare coding variant in the zinc finger ZZ-type and EF-hand domain containing 1 (*ZZEF1)* (rs148096119; *P* = 1.12E-06) exhibited episodic memory decline (**Supplementary Fig. S13**).

**Figure 3.**
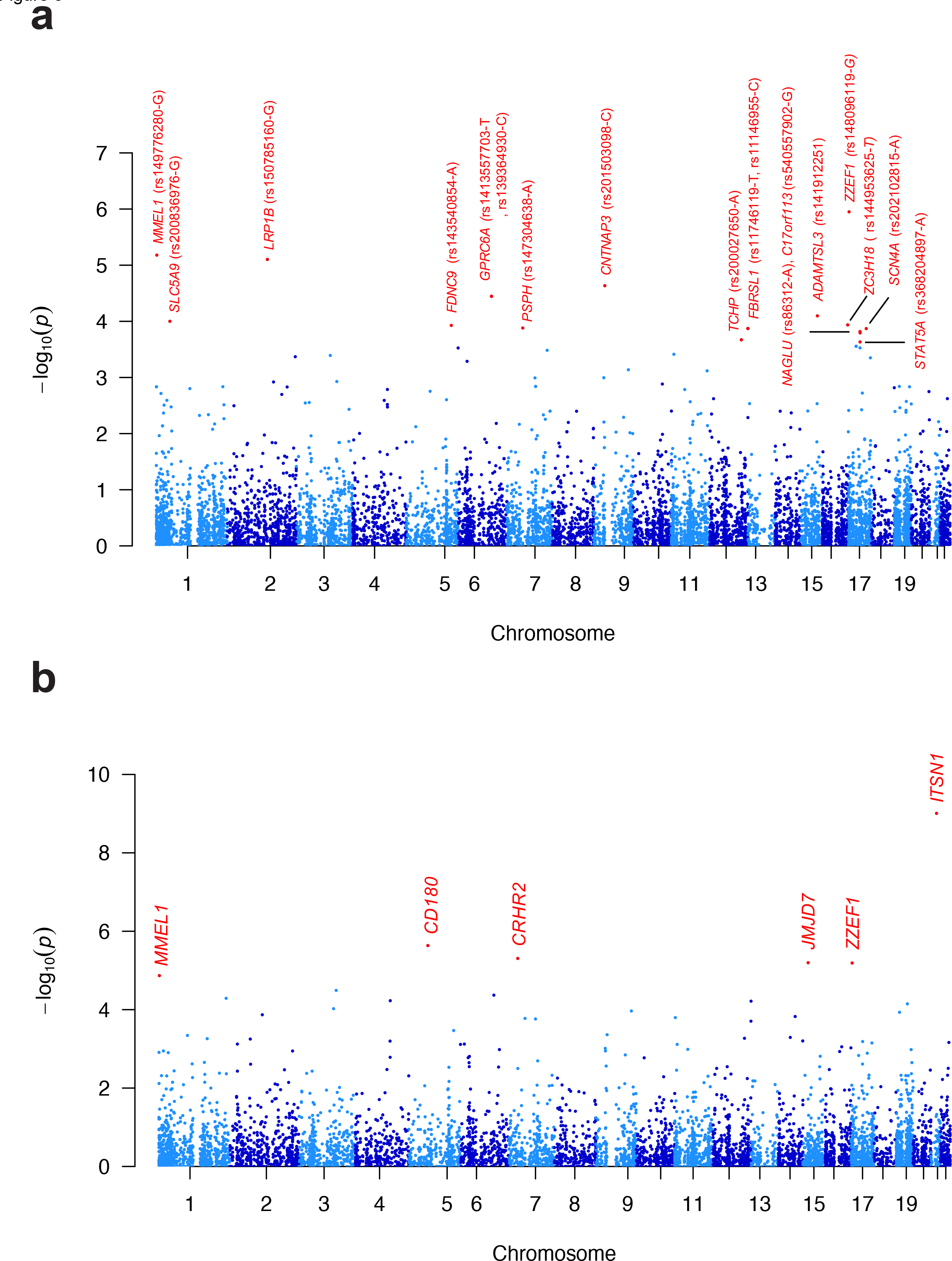
Rare variant association analyses. a. The Manhattan plot for 8,562 prioritized rare coding variants across the exome. See **Supplementary Table S2** for variant annotations. **b.** The Manhattan plot for the gene-based rare variant association analysis.

Next, we conducted a gene-based rare variant association analysis to investigate the association between rare variants and residual memory slope at the gene level. In this analysis, we investigated 10,672 genes containing multiple rare nonsynonymous variants predicted to be functional (CADD score ≥ 20), each with a total occurrence of at least 5 in our study cohort (see **Methods**). For each of these genes, we aggregated its rare nonsynonymous variants and performed a variant-set association test to assess the association of variant sets with residual memory slope. We opted to use SKATO as the variant-set association test as it combines the strengths of the burden test and SKAT. Our analysis revealed significant rare variant associations with six genes at an FDR < 0.05 (**Fig. 3b** and **Supplementary Table S3**). The top gene was intersectin 1 (*ITSN1*) (*P* = 9.83E-10). We further examined rare coding variants in this gene using the burden test and confirmed a significant burden of rare coding variants in this gene that may contribute to episodic memory decline (*P* = 2.43E-06).

Overall, there were 20 risk genes implicated by either single or gene-based rare variant associations (**Table 2**). Gene expression analyses revealed that most of these risk genes were widely expressed in brain tissues (**Supplementary Fig. S14**). Since AD progression is one possible reason underlying episodic memory decline, we examined AD genes previously implicated by rare coding variants (*APP*, *PSEN1*, *PSEN2*, and *TREM2*)^43^ but didn’t observe significant association signals. However, two genes playing protective roles in AD pathology via regulation of amyloid-β level (*LRP1B* and *MMEL1*)^44,45^ were implicated by single rare variant association, (rs150785160-G and rs149776280-G, respectively). We examined these two rare coding variants in the ADSP study cohort. Although carriers of rs149776280-G were not found in the ADSP cohort, we confirmed that the carriers of rs150785160-G have a higher risk of developing AD (Odds ratio = 5.7; *P* = 0.035).

### Replication study

We utilized 1,680 subjects of EUR ancestry without an AD diagnosis from the ROSMAP cohort who had their first visit at or after 65 years old and a minimum of 4 annual episodic memory measurements. Using the same approach as in the LonGenity cohort, we calculated the residual memory slope. The result from the linear mixed model analysis indicated a significant negative correlation between years of follow-up and memory scores (*P* < 2E-16). Older age at the first visit, fewer years of education, and male sex were associated with lower memory scores (*P* < 2E-16, *P* = 1.07E-14, and *P* = 4.41E-14, respectively). The ROSMAP cohort included 526 participants with both the residual memory slope and WGS data available.

For the six genes implicated by gene-based rare variant association, we first examined functional rare coding variants (CADD scores ≥ 20) with MAF < 1% in each gene. However, SKATO did not identify any significant aggregate association. We hypothesized that AJs might have a higher frequency of certain rare causal variants due to the founder effect; certain rare causal variants may occur at lower frequency in the more heterogeneous EUR population. Shifting the focus to functional singleton rare coding variants (CADD scores ≥ 20) in ROSMAP WGS data covered by the initial criteria, we found a significant negative association between the variant burden and residual memory slope in *ITSN1* (*P* = 0.038) and *CRHR2* (*P* = 0.03). At the single variant level, we did not replicate our findings, as most of the putative risk single variants identified in our study cohort either did not appear in the replication cohort or were extremely rare, making our replication cohort likely underpowered to detect their effects. Out of 20 rare coding variants, only two appeared in the replication cohort. Among the three common variants in our cohort, two were rare (MAF < 1%) in the replication cohort. Conversely, by considering AD PRS that characterized aggregate effects of common variants, we replicated and confirmed our finding that AD common polygenic risk significantly contributed to episodic memory decline (adjusted *P* = 2.0E-04), even among individuals without a diagnosis of AD (**Supplementary Fig. S15**).

### Impact of rare variants on protein structure and function

In order to establish additional support for the functional impact of the 25 missense putative risk coding variants implicated by single and gene-based association (**Supplementary Table S2** and **S3**), we studied their contribution to protein stability. The effect of each variant was evaluated separately in an implicit solvent model. Results indicated that most missense variants made the protein (mutant protein) unstable when compared to the wild type (**Supplementary Fig. S16**). Among all, ZZEF1:p.Leu1959Pro and CRHR2:p.Arg148Trp produced the highest instability with a Gibbs free energy change value (ΔG) of 36.79 and 31.94 kcal/mol, respectively (**Supplementary Table S4**). Several of the variants were noted to increase protein stability.

To ascertain the role of missense variants in perturbing the proteins’ overall architecture as well as PPI network, we examined the impact of rare missense variants significantly associated with residual memory slope at amino acid resolution using different protein engineering approaches (**Supplementary Table S5**). We constructed a pool of three-dimensional (3D) structures of proteins that harbor missense variants by using either the experimentally validated crystal structures from PDB or models generated by AlphaFold^33^. Homology modeling was carried out for low-quality structures predicted by AlphaFold using Schrödinger Prime 2022-2 (Prime, Schrödinger, LLC, New York, NY, 2022).

We first focused on variants that altered the secondary structures of the respective proteins. The majority of disease-causing mutations—about 80% of them—have been found to reside in secondary structures^46^. Most of them disturbed the overall configuration of respective proteins, compared to the wild type (**Supplementary Fig. S17-S25**). For instance, contactin-associated protein-like 3 (*CNTNAP3*) variant p.Glu999Gly was located in a conserved EGF-like 2 domain. The wild type Glu999 residue was found to make hydrogen bonds with Ser1001, Ser1169, and Cys1203. However, the Gly999 variant was only able to form a hydrogen bond with Ser1001 (**Supplementary Fig. S25**). Thus, in the case of CNTNAP3:p.Glu999Gly, the combination of the extra length of sidechains as well as polar functional groups allows the wild type Glu999 to make more interactions compared to a non-polar mutant Gly999. Therefore, the Gly999 variant likely destabilized this region as evidenced by the substantial reduction in the number of interactions in CNTNAP3 and confirmed by the stability analysis (**Supplementary Fig. S16**), possibly contributing to episodic memory decline.

### PPI-perturbing alleles in memory impairment

Variants located at the protein interfaces produce significant perturbations and have been shown to be associated with disease phenotypes^47^. Interestingly, we identified 3 rare coding variants that were located at the protein interface where a binding partner (another protein) known to be involved in the pathology of memory-related disorders binds (**Supplementary Table S5**). This indicates that variants found in risk genes in our study likely play a role in memory pathogenesis by interacting with another partner via protein-protein crosstalk.

First, we focused on an interfacial variant (Arg148Trp) found in corticotropin releasing hormone receptor 2 (*CRHR2*). It is a G protein-coupled receptor (GPCR) and has a role in multiple intracellular signaling pathways including adenylate cyclase-cAMP-protein kinase A (PKA), MAPK, and protein kinase C (PKC)^48^. A stable binding of GPCR with its G protein is vital to mediate its signaling. To observe the effect of the Arg148Trp variant, we used the high-resolution 3D structure of CRHR2 bound with G proteins (PDB ID: 6PB1), obtained from PDB^49^. Arg148Trp variant is located at the interface where the Gα subunit binds, thus altering the interactions between CRHR2 and Gα (**Fig. 4** and **Supplementary** Fig. 26). Arg148 formed a hydrogen bond with Gα:Gln390. Interestingly, p.Arg148Trp disrupted this interaction and further amended surface topography due to the change in size, charge and polarity between Arg and Trp (**Fig. 4b and 4c**). To investigate the binding dynamics, stability and energetics of wild type and mutant CRHR2-G protein complexes, all-atom MD simulations were performed. All simulations of the CRHR2-G protein complex maintained their overall structural integrity and a Cα root mean square deviation (RMSD) from the initial structure was under 5 Å (**Supplementary Fig. S27a** and **S27b**). To identify and compare backbone stability and fluctuations of the two complexes, root mean square fluctuation (RMSF) of backbone Cα atoms was analyzed and plotted (**Supplementary Fig. S27c and S27d**). Overall, the backbone of both wild type and mutant CRHR2 complex showed lower fluctuations. Interestingly, residues 27-35 (Gα:36-44), 748-751 (CRHR2:217-220), and 824-833 (CRHR2:293-306) fluctuated more in mutant simulations, compared to wild type ones (**Supplementary** Fig. 27c and 27d).

**Figure 4.**
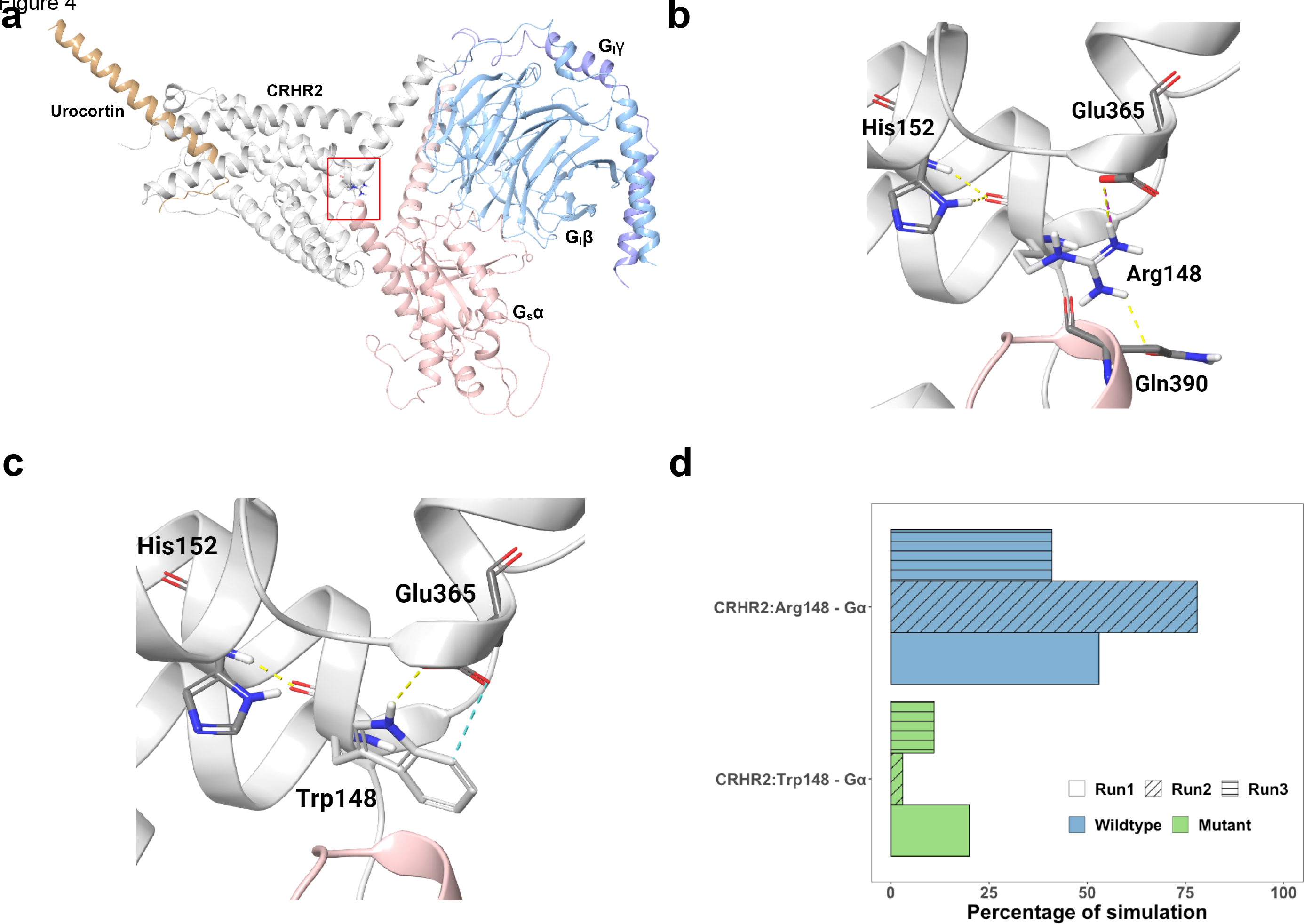
Three-dimensional crystal structure of CRHR2 bound to G proteins. Different subunits are colored differently and labeled. Amino acids are represented with sticks. The red-boxed region in (a) is magnified in the successive images (b and c). (a) 3D structure of CRHR2 complex; (b) Wild type Arg148; (c) Mutant Trp148; (d) The percentage of simulation time during which intermolecular contact was retained between CRHR2:Arg148/Trp148 and Gα protein.

CRHR2:Arg148-Gα protein subunit formed a sustained interaction in all simulations. Such an interaction (CRHR2:Trp148-Gα), however, was found to be very weak in the mutant simulations (**Fig. 4d**). Importantly, wild type Arg148 stabilized the conformation of CRHR2 in such a way that the N-terminal residues Arg38 and His 41 of Gα formed sustained interactions with CRHR2 residues Tyr217 and Glu220 (**Supplementary Figs. S26** and **S28**). However, Gα N-terminal bound less stably with mutant CRHR2. To measure energetic contributions, the free energy of binding (ΔGbind) of the Gα subunit to CRHR2 was estimated from frames of MD simulations every 100ns using molecular mechanics-generalized Born surface area (MM-GBSA) method. All three wild type CRHR2-Gα bound simulations showed ΔGbind values (−97.3 ± 7.4 kcal/mol, −78.6 ± 8.8 kcal/mol, and −106.2 ± 13.6 kcal/mol) that were substantially higher than mutant CRHR2-Gα bound simulations (−65.29 ± 17.2 kcal/mol, −70.8 ± 21.1 kcal/mol, and −82.9 ± 17.62 kcal/mol), indicative of stronger binding. This indicates Arg148Trp variant will likely alter CRHR2 downstream signaling due to impaired binding of the Gα subunit to the CRHR2 receptor. These results support previous findings where diminished activity of CRHR2 has been associated with learning, memory, and AD ^50–52^.

Next, we focused on another interfacial variant (p.Thr152Ile) observed in Phosphoserine phosphatase (*PSPH*). Human PSPH is a critical enzyme in the phosphorylated pathway of L-serine biosynthesis. It is formed through the dephosphorylation of phosphoserine via PSPH. The impact of this variant on protein structure and function was evaluated by retrieving 3D coordinates of PSPH bound with phosphoserine substrate from PDB (PDB ID: 1L8L) ^53^. Thr152Ile variant is located near the vicinity of the binding site of PSPH where its natural substrate, phosphoserine binds (**Fig. 5a**). Thr152 formed two hydrogen bonds with Phe58 and Ser109. However, p.Thr152Ile was only able to form a hydrogen bond with Phe58 (**Fig. 5b** and **5c**). To investigate the influence of this variant on binding mode, interaction stability, and binding affinity of phosphoserine for PSPH, we ran all-atoms MD simulations in triplicates. All simulations remained stabilized throughout the simulation runs and a Cα RMSD from the initial structure was under 5 Å (**Supplementary Fig. S29a** and **S29b**). Mutant PSPH simulations showed more fluctuations especially residues 193-206, compared to wild type runs (**Supplementary Fig. S29c** and **S29d**). Importantly, residues (Asp20, Asp22, Ser23, Glu29, and Arg202) that were part of the PSPH active binding site also fluctuated more in mutant simulations compared to wild type. Interestingly, phosphoserine maintained more sustained interactions with essential binding residues of wild type PSPH, compared to mutant PSPH (**Fig. 5d, 5d** and **Supplementary Fig. S30**). Previous mutagenesis studies focused on active binding site residues of PSPH showed that point mutations of most of these residues significantly diminished enzymatic activity, likely via altered substrate binding^53^. We also computed the binding affinity (ΔGbind) of phosphoserine to PSPH in each simulation run after every 100ns. Wild type simulations demonstrated ΔGbind values (−31.7 ± 3.8 kcal/mol, −29.6 ± 2.9 kcal/mol, and −36.3 ± 3.6 kcal/mol) that were higher than mutant simulations (−24.7 ± 2.8 kcal/mol, −17.4 ± 10.5 kcal/mol, and −29.1 ± 3.5 kcal/mol) This indicates that Thr152Ile likely diminished PSPH enzymatic activity via altering phosphoserine binding to phosphoserine phosphatase. Our findings are in agreement with previous results that L-serine protects hippocampal neurons from oxidative stress-mediated mitochondrial damage and apoptotic cell death in a mouse model^54^. Additionally, reduced production of L-serine via PSPH as well as diminished activities of phosphoprotein phosphatase has been associated with cognitive decline in AD^55,56^.

**Figure 5.**
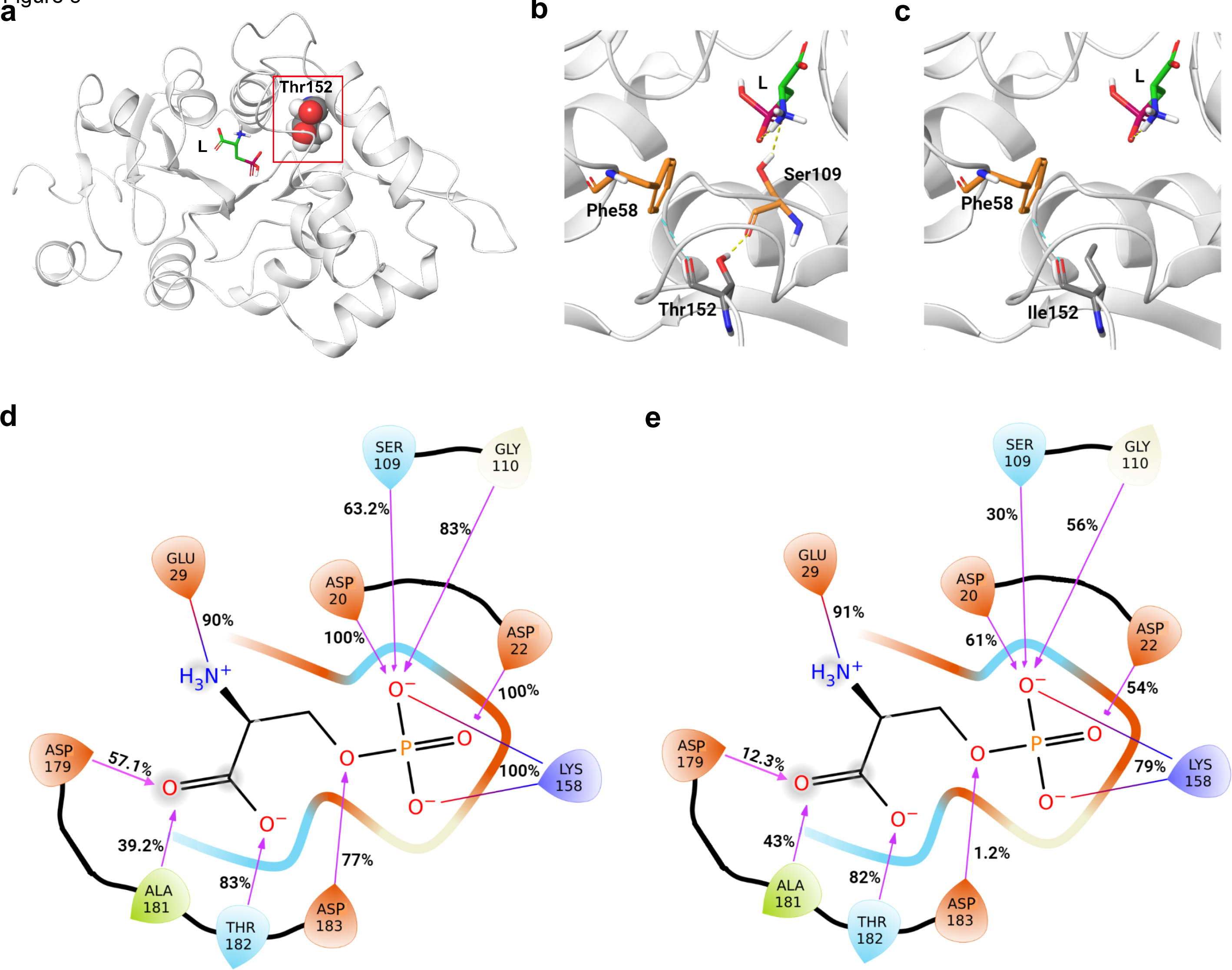
Three-dimensional structure of PSPH bound phosphoserine. Amino acids are represented with sticks. The red-boxed region in (a) is magnified in the successive images (b and c). (a) Crystal structure of CRHR2 complex; (b) Wild type Thr152; (c) Mutant Ile152; (d). Average percentage of simulation time a wild type PSPH residue maintains contact with phosphoserine ligand (L); (e) Average percentage of simulation time a mutant PSPH residue maintains contact with phosphoserine. Charged and polar amino acids are shown in orange and blue color, respectively.

## DISCUSSION

In this study, we conducted a comprehensive genetic analysis of episodic memory in older adults from a longitudinal cohort, LonGenity, to elucidate the contributions of both common and rare coding variants. Our investigation revealed robust aggregate effects of both common and rare variants on episodic memory decline in the LonGenity cohort that were subsequently replicated in the ROSMAP cohort. PRS analysis and gene-based rare variant association analysis underscored the significant contributions of common polygenic risk of AD and rare coding variants in *ITSN1* and *CRHR2*, respectively, to episodic memory decline. Single-variant association analysis identified both putative risk common and rare coding variants in genes previously associated with learning and memory. Notably, the putative risk rare coding variants exhibited ∼1.5 times larger effect sizes than the identified common variants, suggesting an important role of rare variants in the pathological decline of episodic memory. To establish the functional impact of these identified risk rare coding variants, we applied protein modeling and all-atom MD simulations, revealing two novel genetic connections, *CRHR2* and *PSPH*, to memory pathologies.

We demonstrated that the common polygenetic risk of AD contributed to episodic memory decline, a finding that may not be surprising in a cohort that included participants with AD. However, even after excluding a small number of LonGenity participants who either had or were at high risk for developing dementia (subjects with memory impairment and cognitive or functional decline during their follow-up period), the association between AD PRS and episodic memory decline remained significant. This observation persisted in the ROSMAP replication cohort that excluded participants with AD diagnosis. We propose two potential explanations for these findings. First, a number of participants in both cohorts may have had early pre-clinical stages of AD pathology. Second, there may be a genetic overlap between AD and episodic memory decline contributing to the observed association. Despite this, AD common polygenetic risk only accounted for 2∼3% of the variance in residual memory slope in both discovery and replication cohorts. This contribution may be expected to be more substantial in a cohort where AD progression is the major factor underlying episodic memory decline. Nevertheless, acknowledging the contribution of AD PRS to age-related episodic memory decline, irrespective of AD diagnoses, may aid in risk prediction and identification of underlying biological causes.

We identified a total of 24 rare variant associations for episodic memory decline, 18 at the variant and 6 at the gene level. Replicating single rare variant associations was often unfeasible due to their low occurrence, given the limited sample size of the ROSMAP replication study cohort (*n* = 526). However, using the ADSP cohort with a substantially larger sample size (*n* = 9,976), we confirmed the association of AD risk with the putative risk rare coding variant in *LRP1B*, which has been previously shown to protect against AD pathology^44^. Among the six putative risk genes implicated by the aggregate effects of rare coding variants, we replicated two gene-based associations (*ITSN1* and *CRHR2*). The limited sample size, coupled with the high genetic heterogeneity of episodic memory decline, may account for the challenge of achieving more substantial replication. Overall, we identified three robust rare-variant associations, supported by replications in independent cohorts, that implicated distinct pathogenic pathways in episodic memory decline – AD (*LRP1B*), hippocampal plasticity (*ITSN1*) and corticotropin signaling hormone (*CRHR2*) (**Table 2**). Examining other putative risk genes implicated by rare variants revealed further evidence of hippocampal pathologies. Similar to *ITSN1*, *ZZEF1* also contributed to the modulation of hippocampal plasticity, and their disruption caused memory impairment in mouse knockout models^57^. The CNTNAP3 protein has been found to be highly expressed in the cortex and hippocampus of the mouse postnatal brain at P7 and P14^58^. Moreover, male Cntnap3-null mice demonstrated deficits in spatial learning and prominent repetitive behaviors^59^. Additionally, there are other putative risk genes for episodic memory impairment supported by mouse knock-out models through different mechanisms, such as hyperphosphorylated tau (*NAGLU*)^60^, and brain *STAT5A* signaling^61^.

The hippocampus is crucial for learning and memory, harboring neural stem cells (NSCs) with a natural capacity to generate new neurons^62,63^. Studies in mammals have consistently shown a decline in hippocampal neurogenesis during physiological aging^64–66^. Like mammals, a decrease in hippocampal neurogenesis has also been observed in human aging^67^. A study focused on human brain tissue revealed diminished angiogenesis, neuroplasticity, and a smaller quiescent pool in the anterior dentate gyrus (DG) when compared to younger tissue^68^. However, the exact molecular mechanisms underlying this decline remain unknown. Recent studies have shown that the reduction in neurogenesis seen during normal aging is further aggravated in neurodegenerative disorders like AD^67^. Interestingly, we identified rare variants in genes (*ZZEF1*, *ITSN1*, *CNTNAP3, CRHR2, PSPH)* that are known to implicate hippocampal functions^54,57,59,69,70^, indicating their involvement in episodic memory decline in our aging cohort.

We were unable to replicate the findings from prior GWAS of episodic memory, cognitive function, AD-and-related phenotypes^15,71^. This may result from differences in study populations, variations in cognitive tests included in phenotype construction, and small sample sizes. In particular, our study cohort was enriched with offspring (49.1%) of centenarians. It is known that centenarians harbor beneficial variants against aging-associated diseases^28,72^. Having inherited protective genotypes that result in resilience to episodic memory decline in our study cohort may compromise the power of identifying pathogenic risk variants. Nevertheless, earlier studies have found other variants within the three risk loci regions in association with related phenotypes, such as pre-frontal cortex development, age-related cognitive decline, cognitive decline in AD, and education attainment^15,71,73–76^, indicating association of these loci to memory formation and general cognitive ability. Our epigenome fine-mapping results suggested that the three GWAS SNPs were located in transcriptionally active regions (**Fig. 2**). rs35990795 was an eQTL of *CLIC6,* which has been implicated in learning^77^, in the putamen that is important in learning, motor control, language, reward, and other cognitive functioning^78^. rs548640610 was located within the neuron and astrocyte-specific regulatory elements (**Fig. 2e**) and implicated *CHRM3. CHRM3* is a member of the larger family of G protein-coupled receptors, which influences both central and peripheral nervous system processes through interaction with acetylcholine. Mutations in this gene previously have been associated AD^79^.

In recent years, the importance of protein interfaces as hubs for disease-associated variants has become more widely recognized^80–82^. An interface mutation can destabilize the interface, prevent the partner from binding, alter the partner’s binding affinity, or stabilize the overall binding complex^83^. Such variants produce significant perturbations and have been shown to be associated with disease phenotypes^47^. In this study, we identified three variants located at the binding interface of *CRHR2*, *PSPH,* and *CD180* genes. As a proof of concept, *CRHR2* and *PSPH* interfacial variants were systematically studied to uncover their molecular insights using protein modeling and all-atom MD simulations. Results showed that both *CRHR2* and *PSPH* interfacial variants destabilized the binding interface and substantially altered the binding affinity of their respective natural ligands (**Fig. 4** and **5**). Here, our CRHR2 findings are supported by previous studies where they showed that CRHR2 null mice exhibited a phenotype of MCI as well as pathological changes indicative of early AD^50^. Impaired corticotrophin hormone (CRH) signaling via CRHR1 and CRHR2 has also been linked to learning and memory^51,52^. Impaired CRH signaling has been shown to alter hippocampal neurogenesis, spatial memory, and the activity of hippocampal neural stem cells (hiNSCs) under physiological conditions^70^. These compelling results support our finding of a rare interfacial variant CRHR2:p.Arg148Trp that is associated with episodic memory decline. Similarly, PSPH:p.Thr152Ile simulation results demonstrated that phosphoserine bound more stably to wild type PSPH than to mutant PSPH (**Fig. 5**). PSPH:p.Thr152Ile also altered the binding stability of phosphoserine with critical residues of PSPH (**Supplementary Fig. S30**). In a mouse model, the treatment of L-serine demonstrated a protective effect on hippocampal neurons, guarding against oxidative stress-mediated mitochondrial damage and apoptotic cell death^54^. Altogether, our findings indicate that Thr152Ile is likely to diminish PSPH enzymatic activity via altering phosphoserine binding to phosphoserine phosphatase.

While we provided mechanistic insights into two interfacial variants, it should not be inferred that other coding variants do not play a role in memory pathology. We limited the detailed characterization of variants to a subset of interfacial variants to avoid over- interpretation of MD simulation results for which we lacked a convincing protein relevant mechanism. For instance, CD180:p.Gly231 is located at the binding interface of CD180 and MD1. Gly231Arg substantially altered the binding interface of CD180 to MD1 (**Supplementary Fig. S24d-e**). Importantly, *CD180* has been reported as the closest gene to a GWAS signal associated with general cognitive ability^84^. However, its biological relevance to episodic memory decline remains unknown.

We have identified novel associations of both common and rare variants with episodic memory decline; however, future molecular studies would be required to confirm their underlying biological mechanisms. Overall, our findings advance the current understanding of the biology of episodic memory decline and could lead to new drug targets for preventing or treating memory impairment.

## Data Availability

All the data produced in the present study are available upon reasonable request to the authors

## ACKNOWLEDGEMENTS

We are grateful to the individuals for their persistent participation in our longitudinal study of memory decline. We thank Sequencing and Lab Operations team at Regeneron RGC for their execution and oversight of the exome sequencing. We are also thankful for the support from Gregory Klein for his help in accessing RADC data.

## CONSENT STATEMENT

Written informed consent was obtained from all study participants.

