## Supplemental file 1 for "Rare genetic coding variants associated with age-related episodic memory decline implicate distinct memory pathologies in the hippocampus": Supplement.pdf

### Supplementary Note

It is known that skewed phenotype may inflate false positive rates of association especially for rare variants. We applied the conventional log-based transformation to measured memory scores, which substantially mitigated but did not completely remove the skewness of phenotypes (memory scores as well as the slope) (**Supplementary Fig. S2 and S6**). On the other hand, while some extreme transformations may force the transformed phenotype into a normal distribution (e.g. INT) thereby completely removing the skewness of the phenotype, the transformations would not only substantially compromise the power but also distort the natural diversity represented by the phenotype. Therefore, we addressed the issue of skewed phenotype for rare variant association analysis with two steps: first, we applied conventional log-based transformation that reached a balance between mitigating the skewness and preserving the information represented by the phenotype; second, we applied additional procedure to control false positive rates of identified variants that took the skewed phenotype (after transformation) into consideration.

For the second step, the goal was to control the overall false positive rate of identified variants at < 5%. A rare coding variant will be identified if the association P-value (from linear regression) is higher than a threshold (~0.00023 in this study; the highest P-value that passed FDR < 0.05). 594 rare coding variants were identified in our study. For each identified rare coding variant, we further evaluated the chance of identifying false positives that took the skewed phenotype into consideration: For each identified rare variant, we performed a permutation test with 100,000 iterations of randomized residual memory slope, in which the association test P-values were collected to construct the null distribution. The P-values for observed associations below the significant threshold (0.00023) were collected (**Supplementary Fig. S4**). A strict Bonferroni correction for multiple-testing was applied to the randomness tests to ensure that the false-positive rate of identifying each final list of rare variants is under 5%. By focusing on rare non-synonymous variants with CADD scores ≥ 20 that occur in at least 5 carriers, we control the false positives below 5% for significant findings despite the skewness.

**a.**

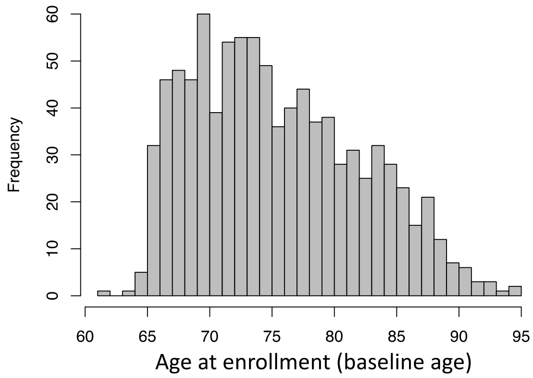


**b.**

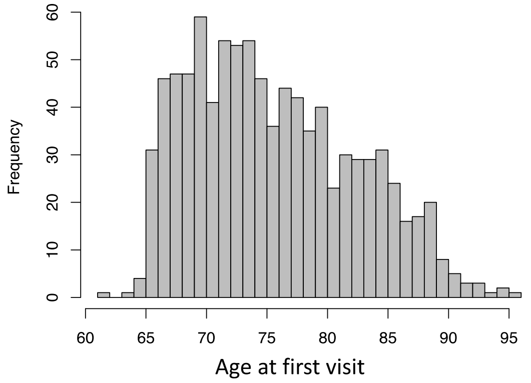


**Supplementary Figure S1. The age distribution of 923 subjects with at least two memory measurements. a.** Age at enrollment (baseline age). **b.** Age at first visit.

**a.**

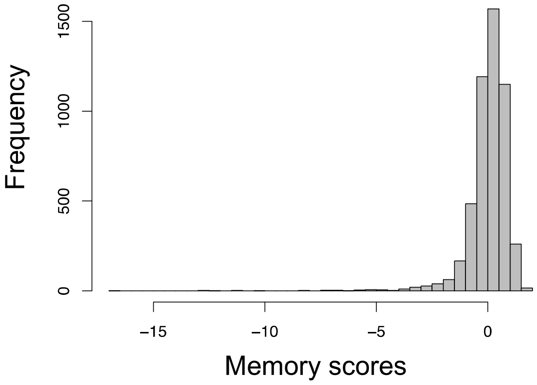


**b.**

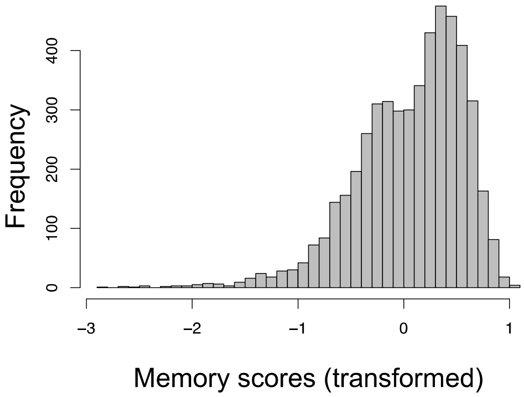


**Supplementary Figure S2. The distribution of measured memory scores in all waves. a.** The original composite memory scores**. b.** The memory scores after transformation.

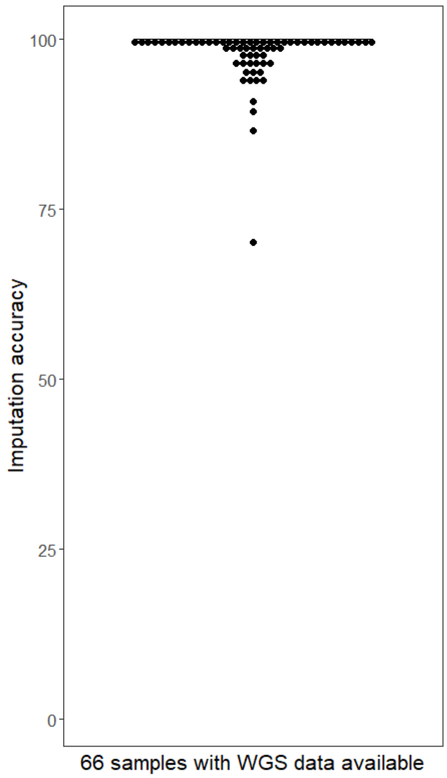


**Supplementary Figure S3. Evaluation of imputation accuracy.** 66 subjects for whom whole genome sequencing data was available were used to evaluate accuracy of imputed genotypes.

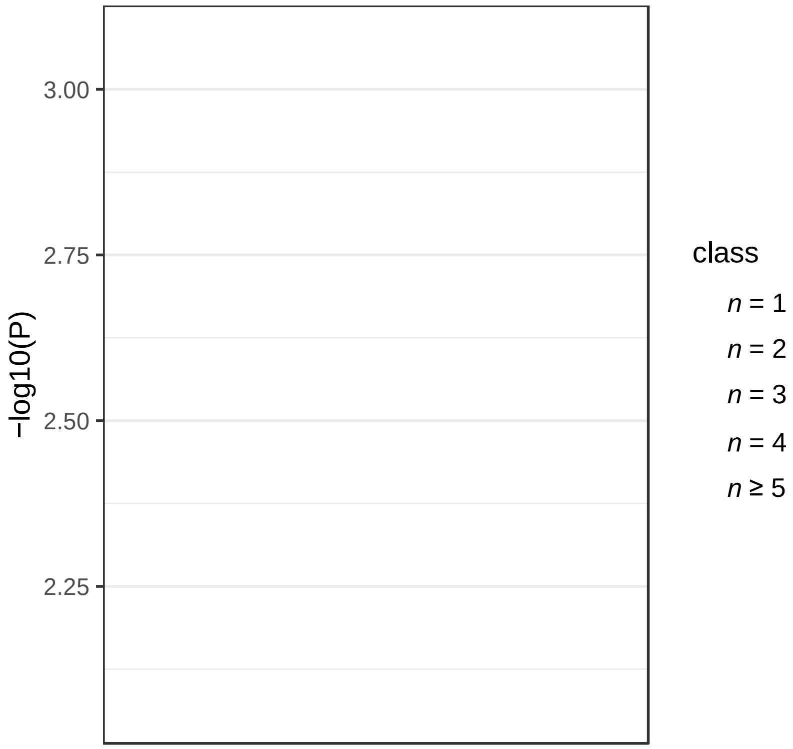


**Supplementary Figure S4. The permutation test to evaluate the false positive rate for identified rare variants.**

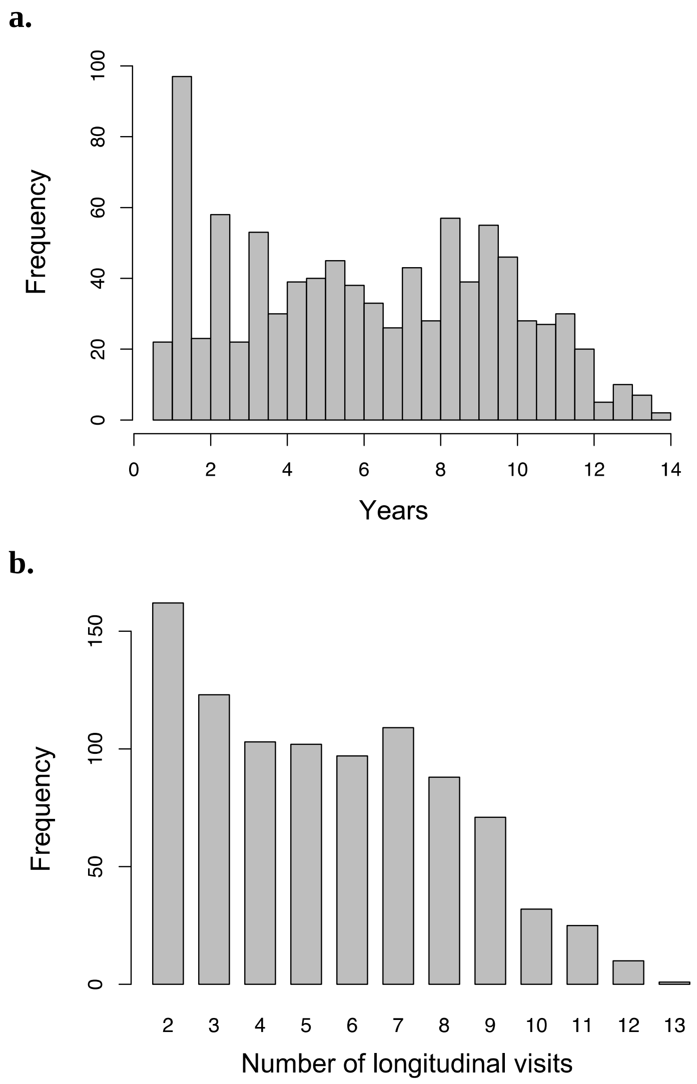


**Supplementary Figure S5. Years of follow-up and number of cognitive assessments per each participant who contributed data to the longitudinal study of memory. a.** Follow-up time in years. **b.** Number of longitudinal cognitive assessments.

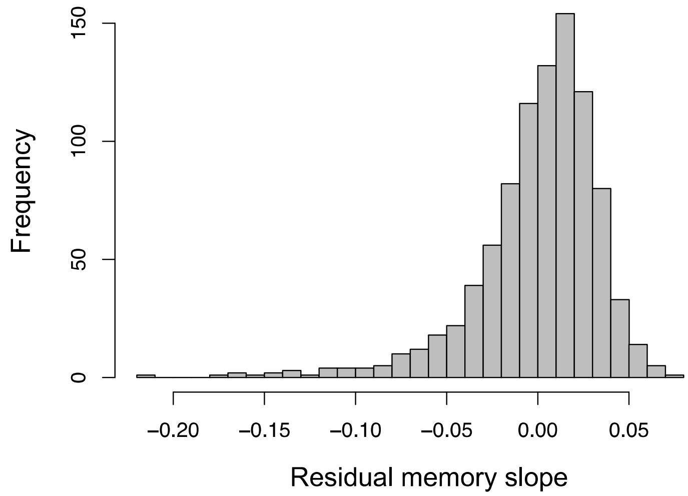


**Supplementary Figure S6. The distribution of residual memory slope for 923 subjects.**

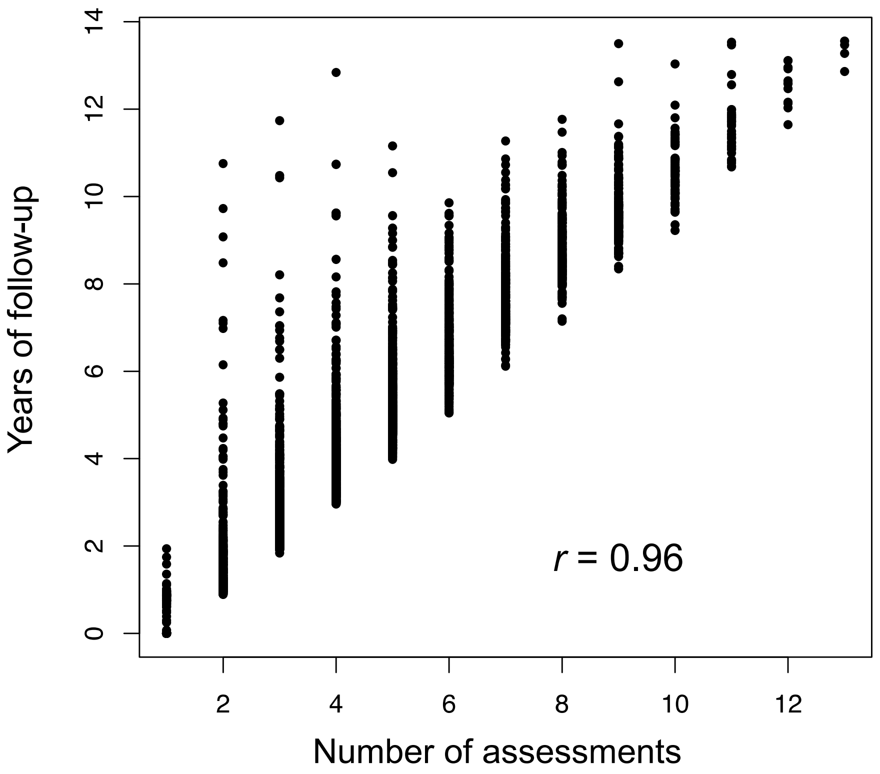


**Supplementary Figure S7. Correlation between number of assessments and follow-up years.**

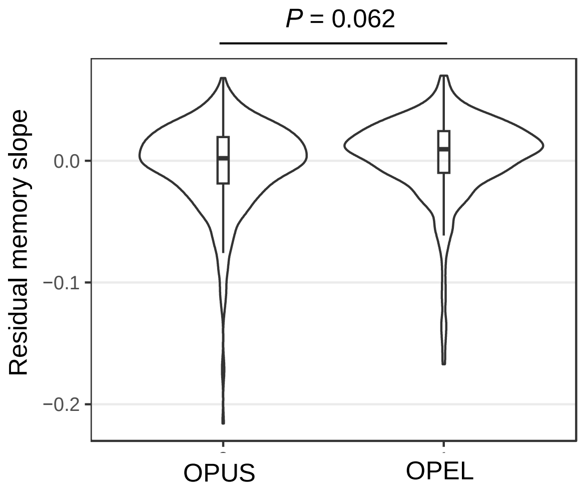


**Supplementary Figure S8. The comparison of the residual memory slope between OPEL and OPUS.** OPEL, offspring of parents with exceptional longevity; OPUS, offspring of parents usual survival.

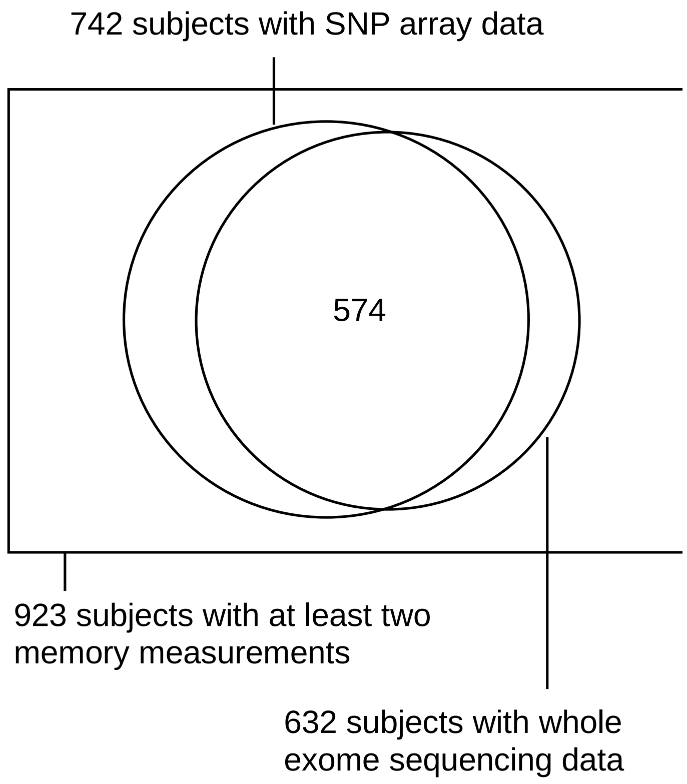


**Supplementary Figure S9. The Venn diagram of discovery study cohorts for episodic memory decline (*n* = 923), common variant association (*n* = 742) and rare variant association (*n* = 632)**

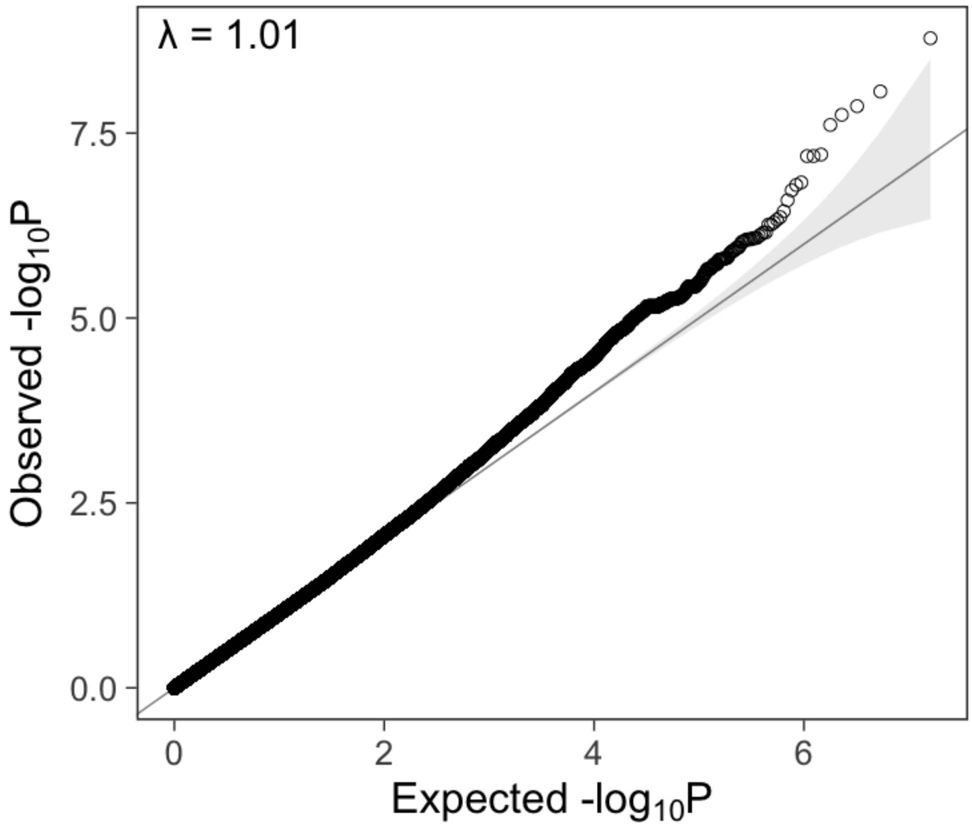


**Supplementary Figure S10. The QQ plot for common variants (MAF ≥ 1%) in the GWAS.**

**a.**

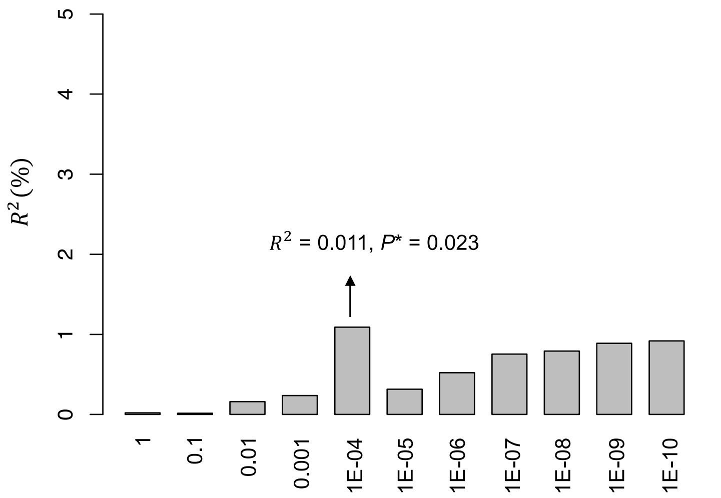


**b.**

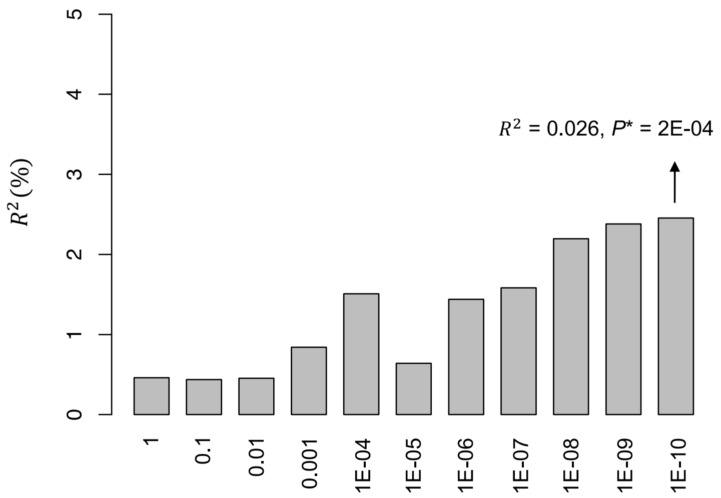


**Supplementary Figure S11. AD PRS contributes to episodic memory decline in the LonGenity discovery cohort.** The X-axis shows the *P*-value thresholds applied in the PRS analysis using PRSice-2. $P^*$ denotes an adjusted *P*-value after correction of running 11 tests with different *P*-value thresholds. LonGenity participants with a higher AD PRS tended to have a lower residual memory slope. Different LD clumping parameters were set to run PRSice-2: **a.** clump-kb = 250kb, clump-r2 = 0.1. **b.** clump-kb = 500kb, clump-r2 = 0.

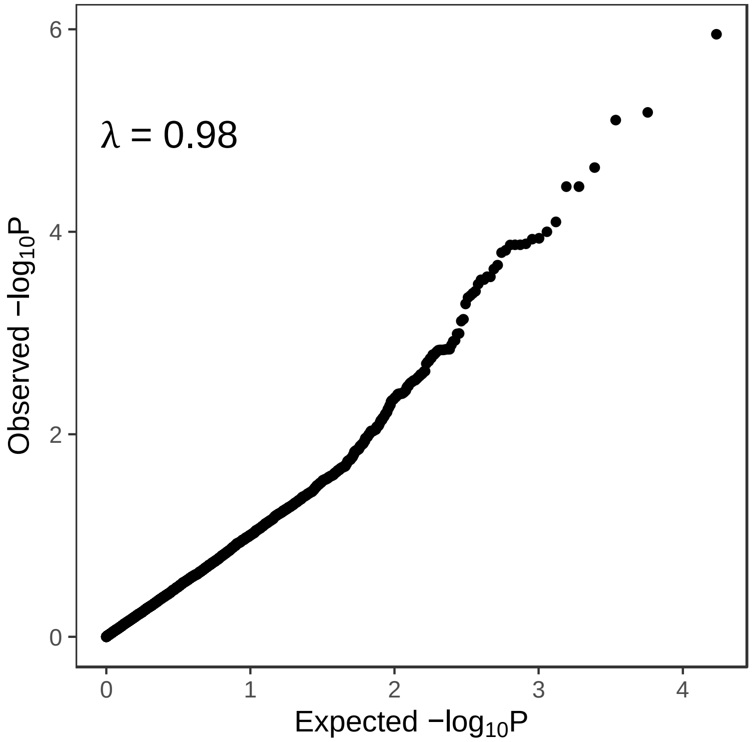


**Supplementary Figure S12. The QQ plot for the rare coding variants association analysis.**

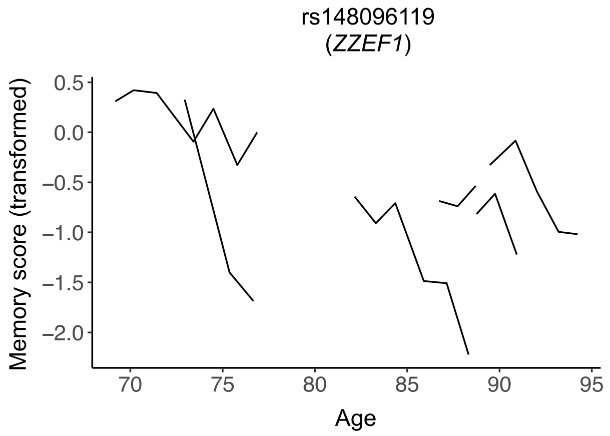


**Supplementary Figure S13. The memory trajectories of carriers for the top prioritized rare coding variant rs148096119.**

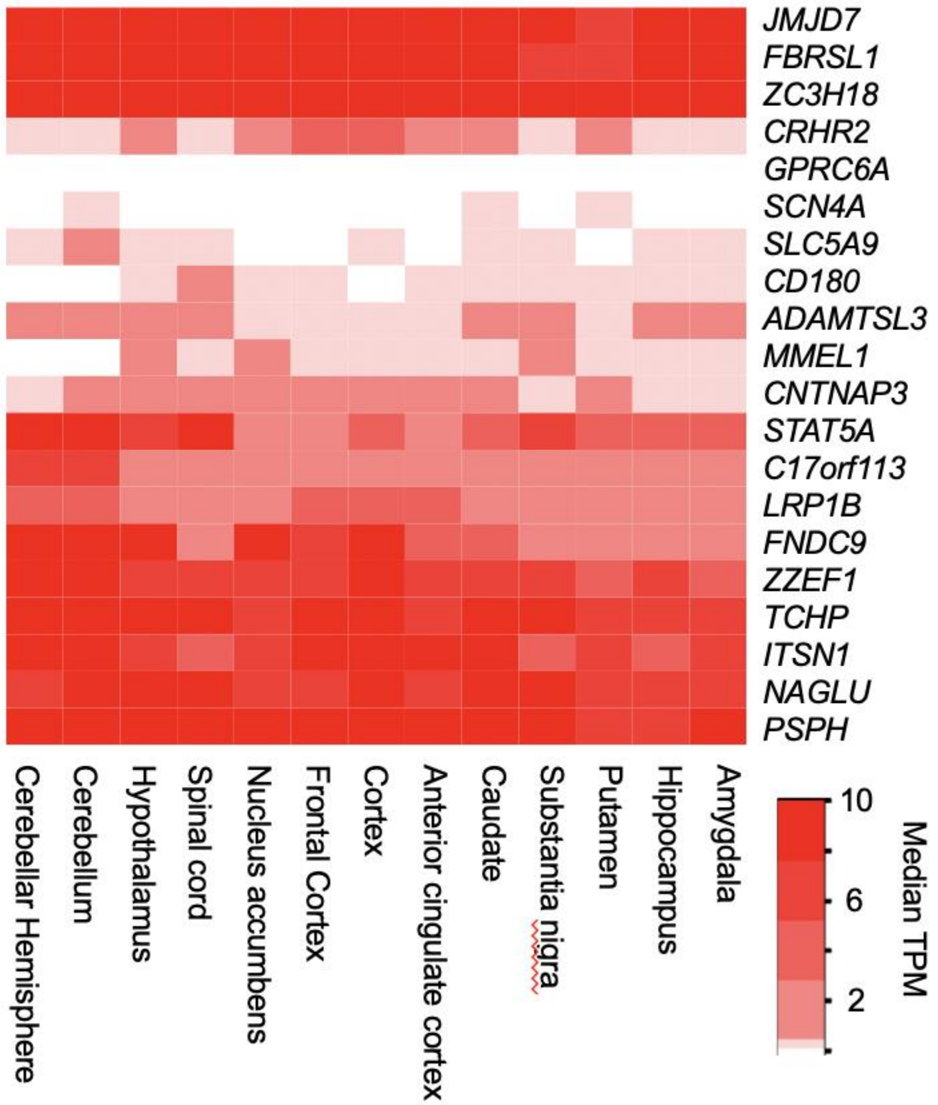


**Supplementary Figure S14. Expression of 20 risk genes across brain tissues.** Expression values of gene transcripts were obtained from subjects age ≥ 65 from GTEX (v.8).

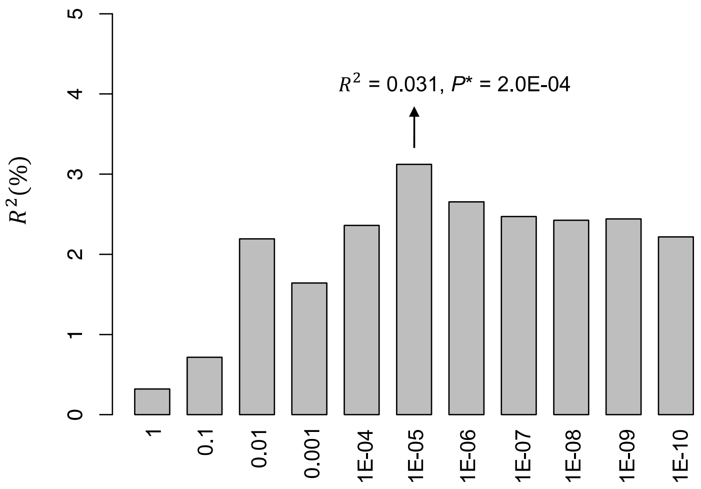


**Supplementary Figure S15. AD PRS contributes to episodic memory decline in the ROSMAP cohort.** The X-axis shows the *P*-value thresholds applied in the PRS analysis using PRSice-2. $P^*$ denotes an adjusted *P*-value after correction of running 11 tests with different *P*-value thresholds. People in the replication cohort with a higher AD PRS tend to have a lower residual memory slope.

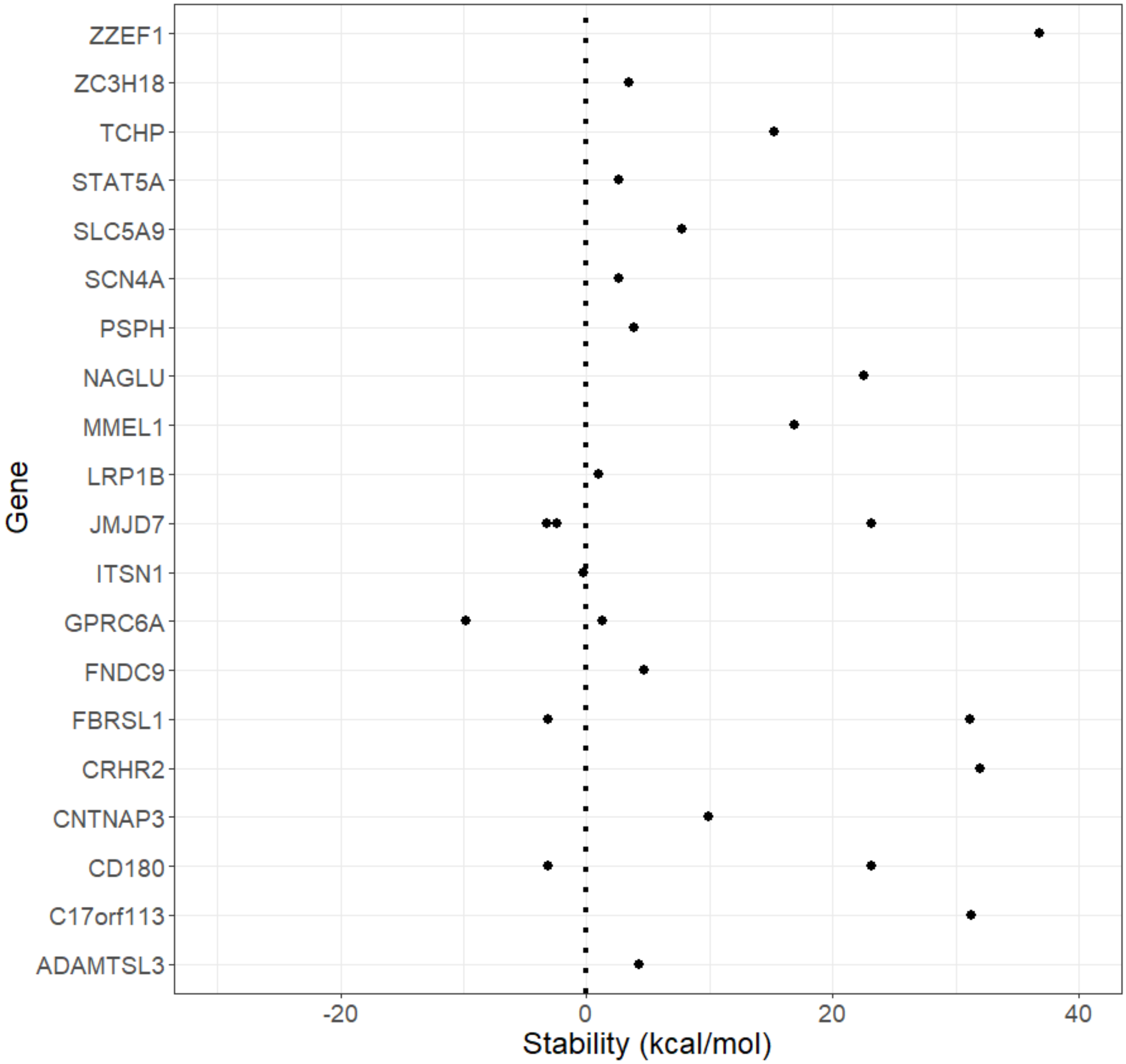


**Supplementary Figure S16. Protein stability analysis of significant rare coding missense variants.** A positive value indicates that the mutant protein is less stable and a negative value indicates that the mutant protein is more stable compared to the wild type protein.

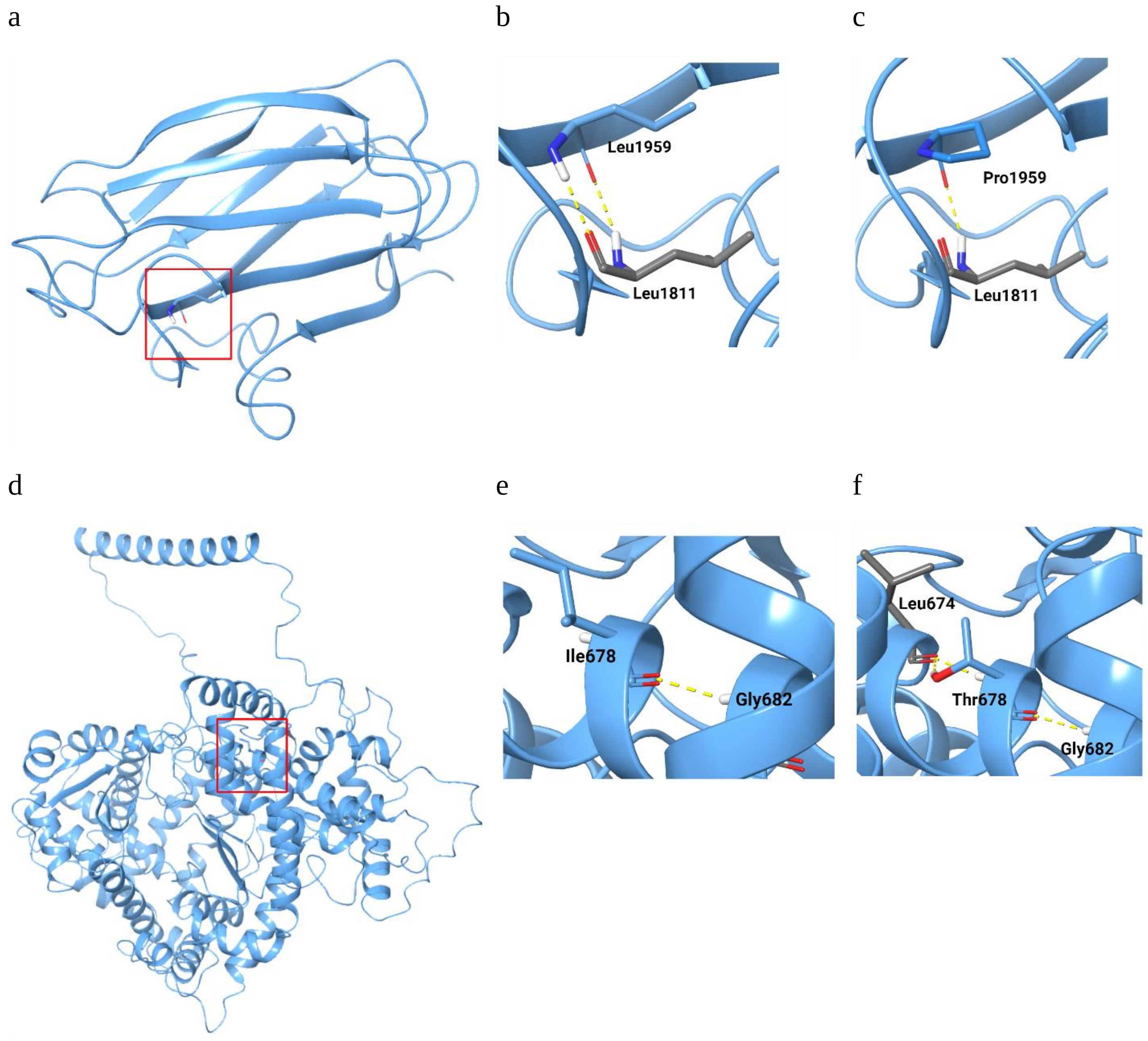


**Supplementary Figure S17. Modeled 3D structure of ZZEF1 and MMEL1.** The proteins are shown in blue cartoon form and amino acids are represented with stick representations. The red-boxed regions shown in (a) and (d) are magnified in successive images. (a) Modeled structure of ZZEF1; (b) Wild type Leu1959; (c) Mutant Pro1959; (d) Modeled structure of MMEL1; (e) Wild type Ile678; (f) Mutant Thr678.

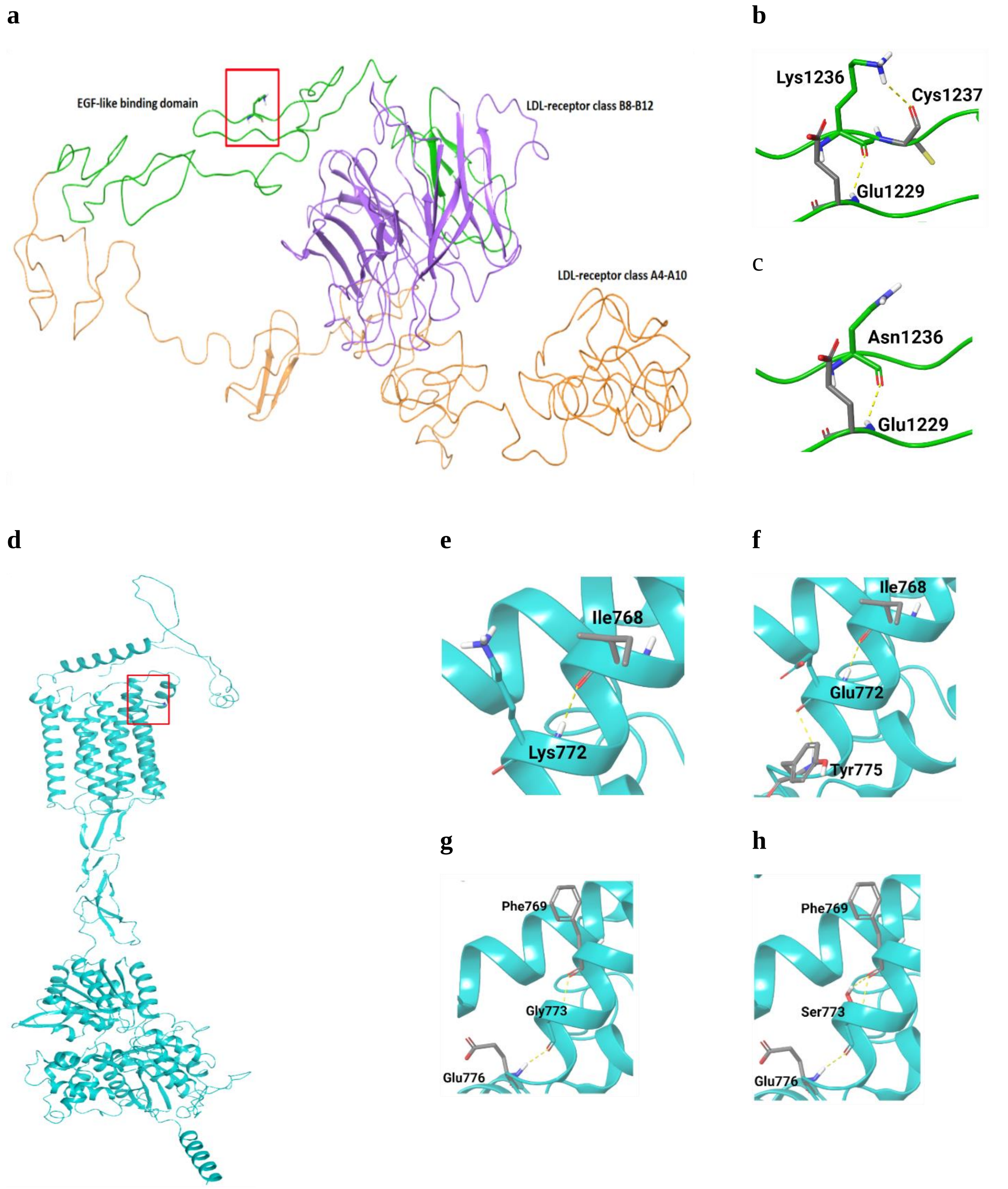


**Supplementary Figure S18. Modeled 3D structure of LRP1B and GPRC6A.** The functional domains of LRP1B protein are shown in different colors and amino acids are represented with stick representations. The red-boxed region shown in (a) and (d) are magnified in successive images. (a) Modeled structure of LRP1B; (b) Wild type Lys1236; (c) Mutant Asn1236; (d) Modeled structure of GPRC6A; (e) Wild type Lys772; (f) Mutant Glu772; (g) Wild type Gly773; (h) Mutant Ser773.

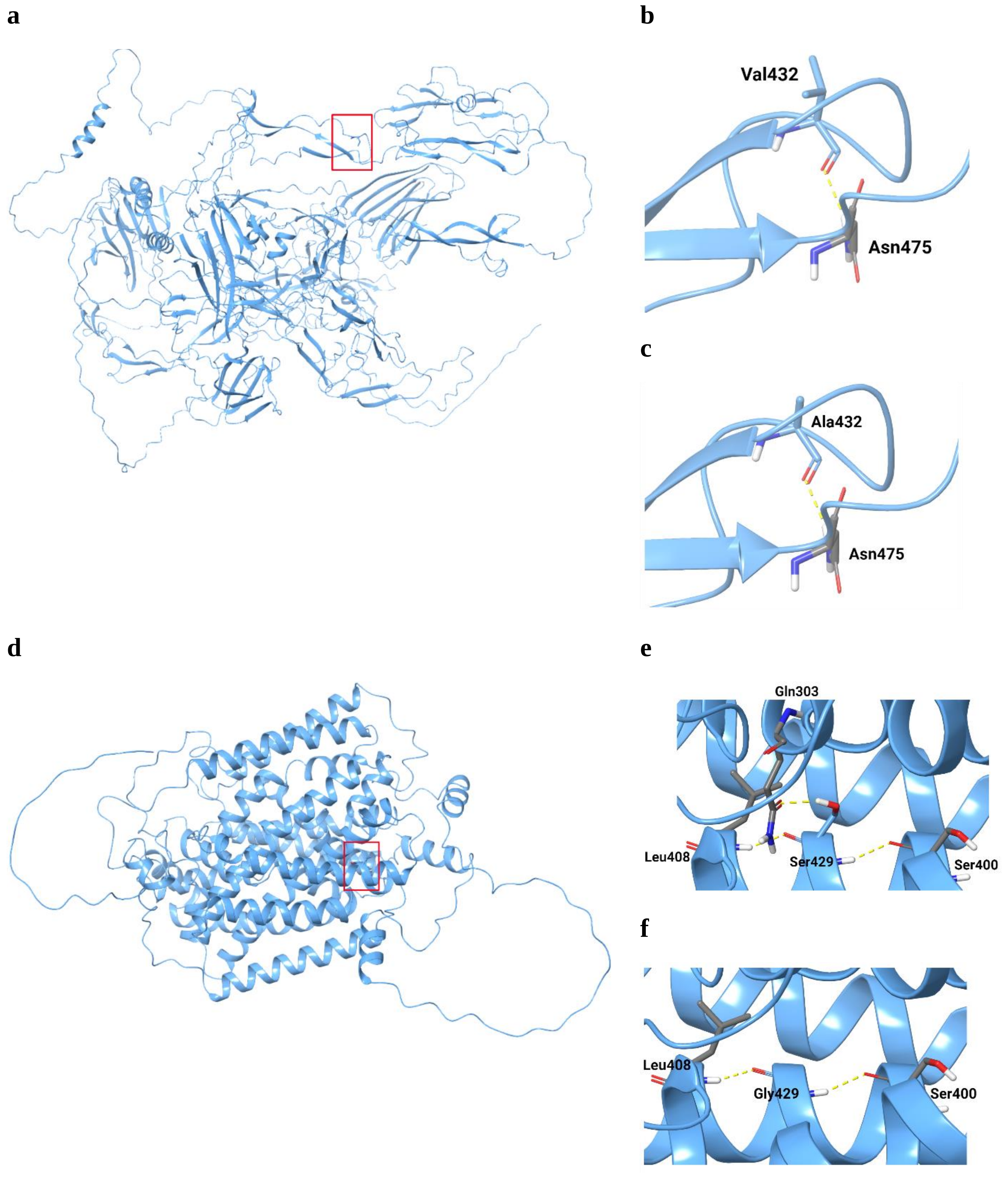


**Supplementary Figure S19. Modeled 3D structure of ADAMTSL3 and SLC5A9.** The proteins are shown in blue cartoon form and amino acids are represented with stick representations. The red-boxed regions shown in (a) and (d) are magnified in successive images. (a) Modeled structure of ADAMTSL3; (b) Wild type Val432; (c) Mutant Ala432; (d) Modeled structure of SLC5A9; (e) Wild type Ser429; (f) Mutant Gly429.

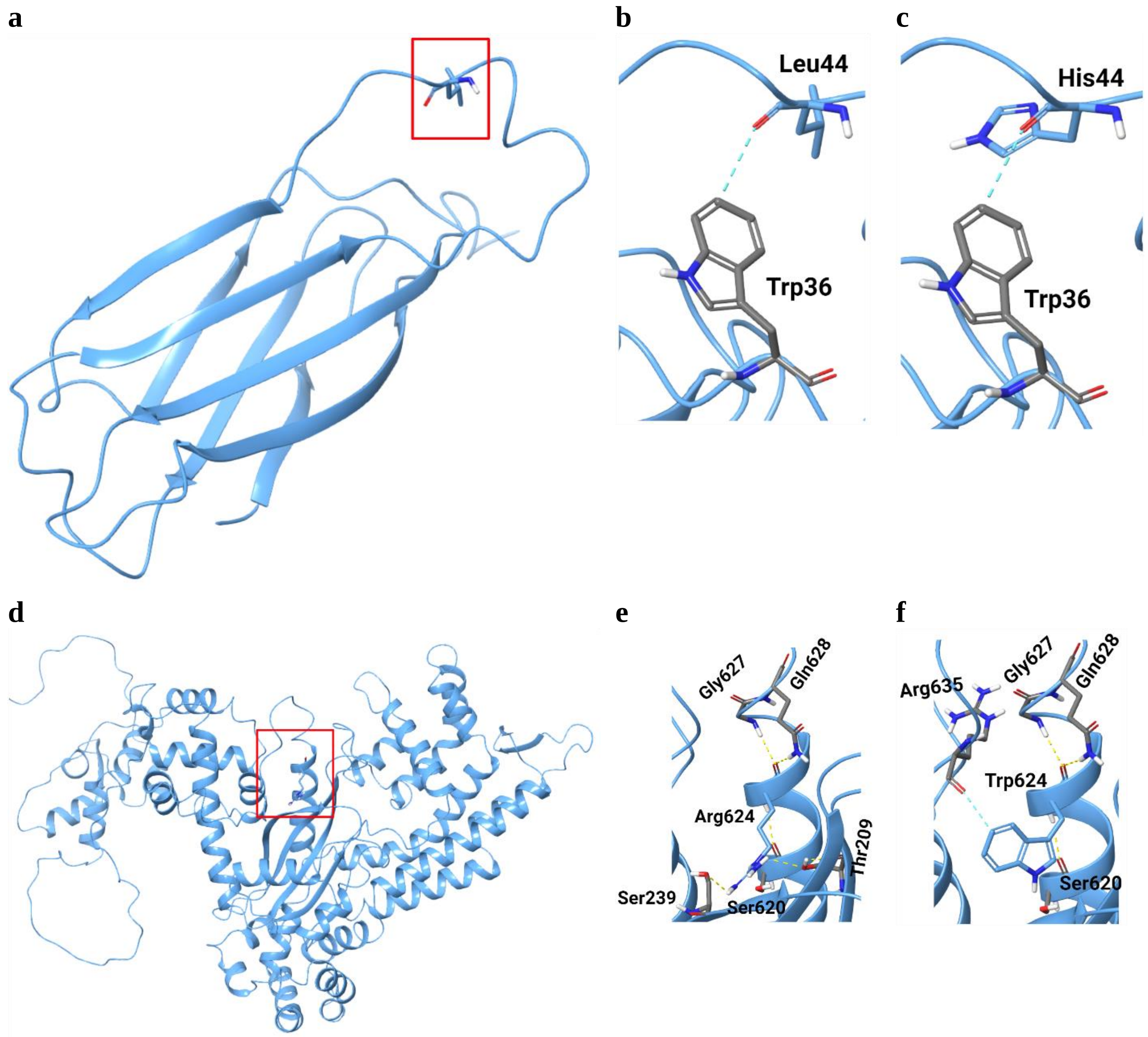


**Supplementary Figure S20. Modeled 3D structure of FNDC9 and C17orf113.** The proteins are shown in blue cartoon form and amino acids are represented with stick representations. The red-boxed regions shown in (a) and (d) are magnified in successive images. (a) Modeled structure of FNDC9; (b) Wild type Leu44; (c) Mutant His44; (d) Modeled structure of C17orf113; (e) Wild type Arg624; (f) Mutant Trp624.

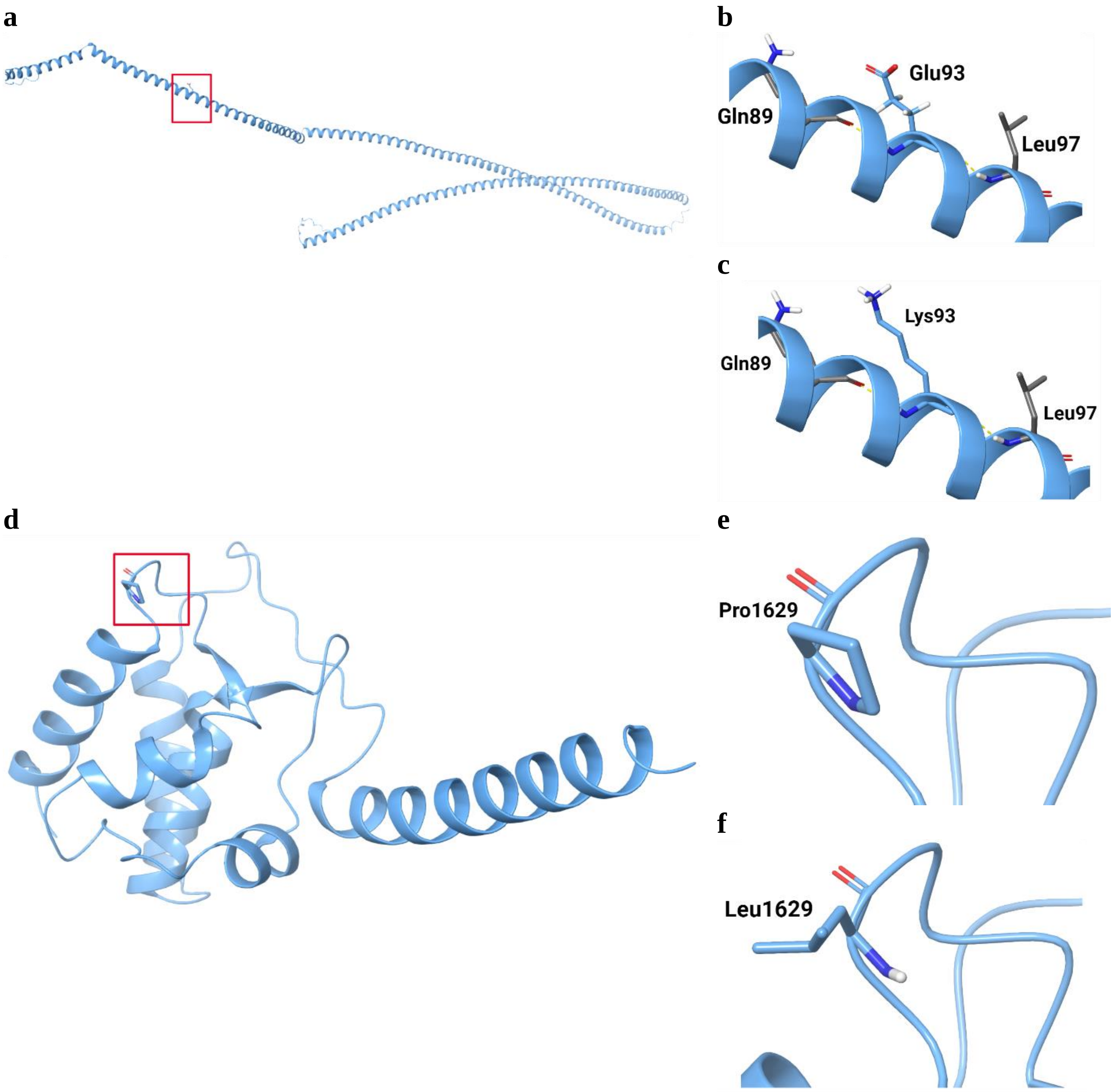


**Supplementary Figure S21. Modeled 3D structure of TCHP and SCN4A.** The proteins are shown in blue cartoon form and amino acids are represented with stick representations. The red-boxed regions shown in (a) and (d) are magnified in successive images. (a) Modeled structure of TCHP; (b) Wild type Glu93; (c) Mutant Lys93; (d) Modeled structure of SCN4A; (e) Wild type Pro1629; (f) Mutant Leu1629.

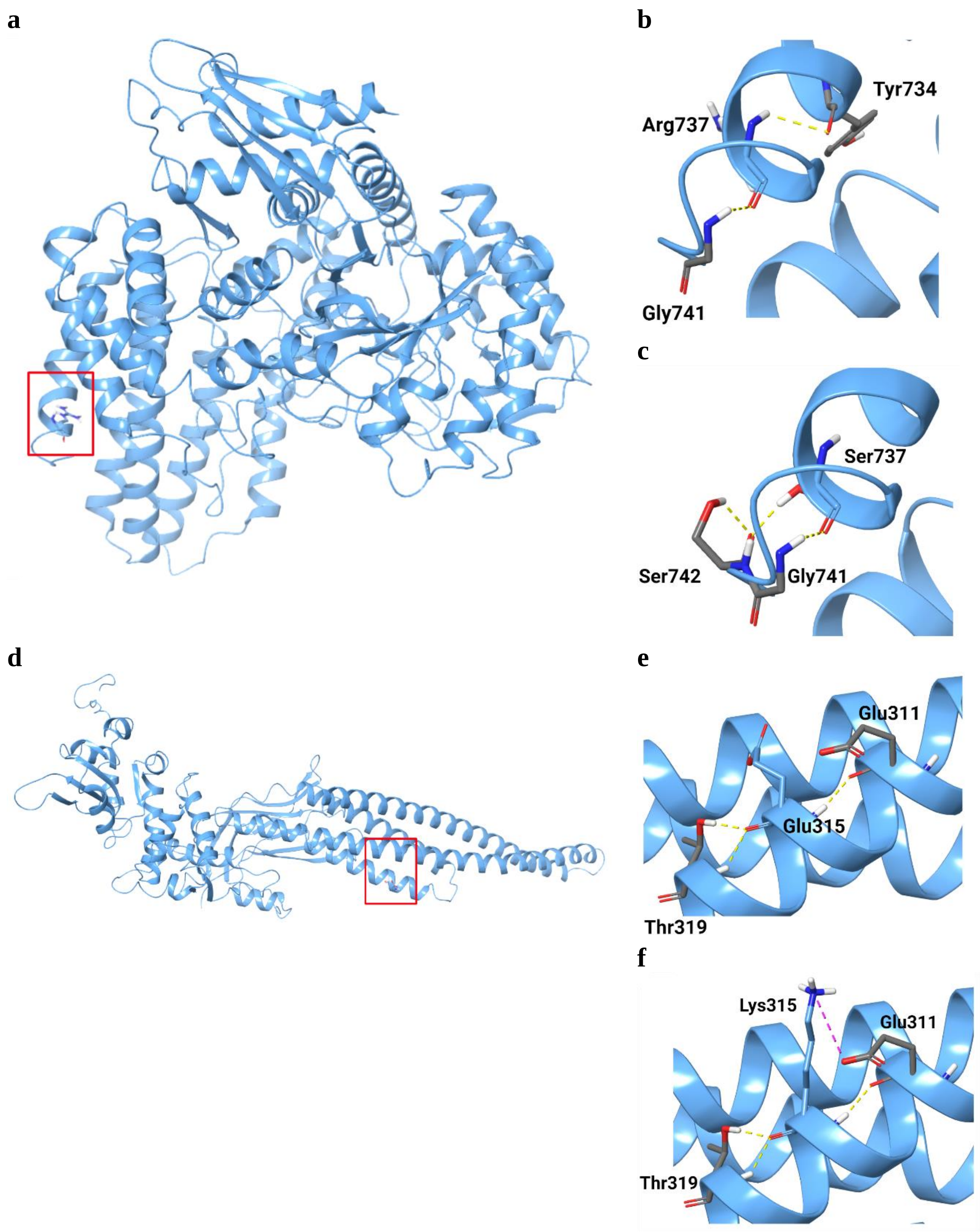


**Supplementary Figure S22. Modeled 3D structure of NAGLU and STAT5A.** The proteins are shown in blue cartoon form and amino acids are represented with stick representations. The red-boxed regions shown in (a) and (d) are magnified in successive images. (a) Modeled structure of NAGLU; (b) Wild type Arg737; (c) Mutant Ser737; (d) Modeled structure of STAT5A; (e) Wild type Glu315; (f) Mutant Lys315.

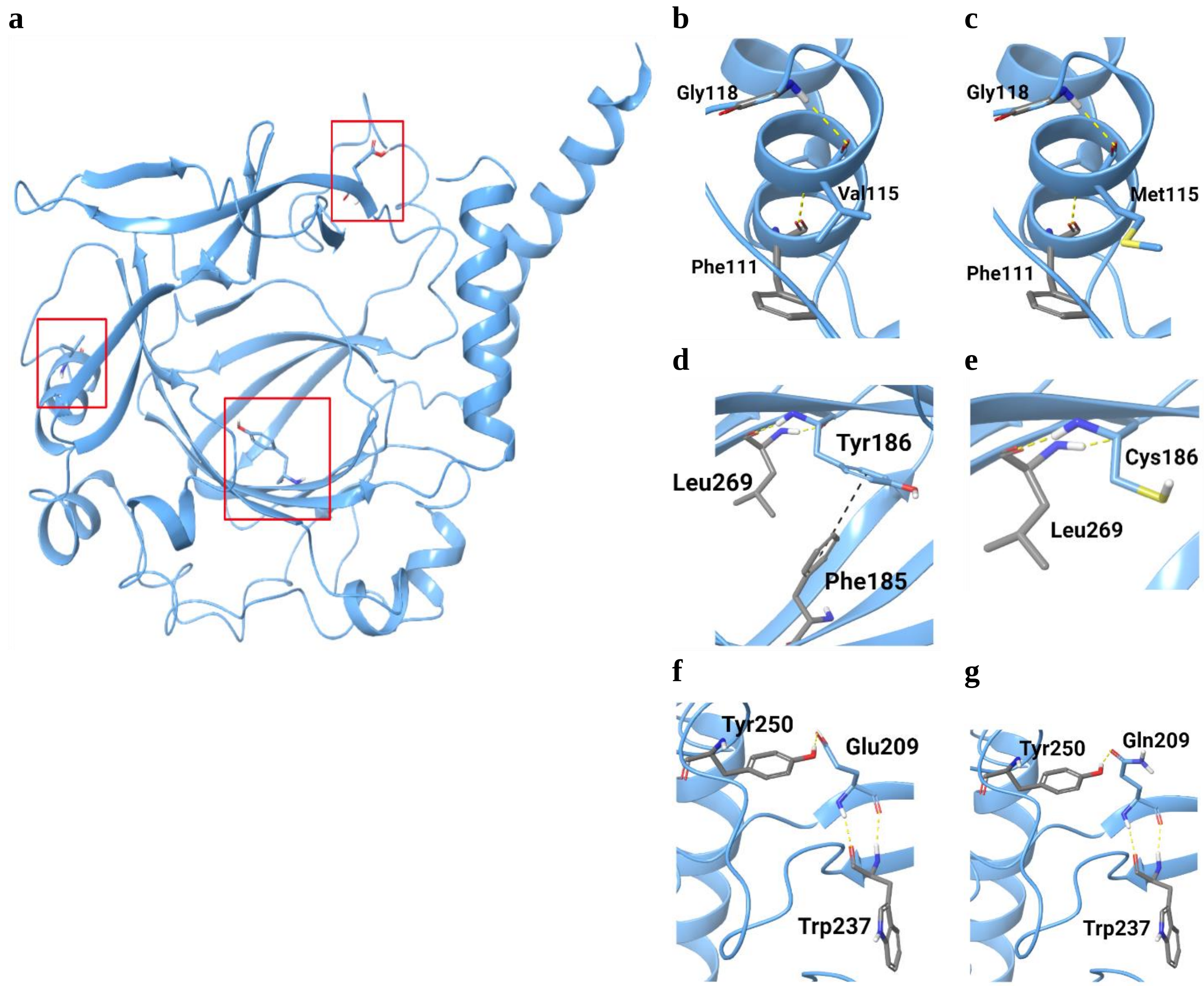


**Supplementary Figure S23. Modeled 3D structure of JMJD7.** The protein is shown in blue cartoon form and amino acids are represented with stick representations. The red-boxed regions shown red in (a) are magnified in successive images. (a) Modeled structure of JMJD7; (b) Wild type Val115; (c) Mutant Met115; (d) Wild type Tyr186; (e) Mutant Cys186; (f) Wild type Glu209; (g) Mutant Gln209.

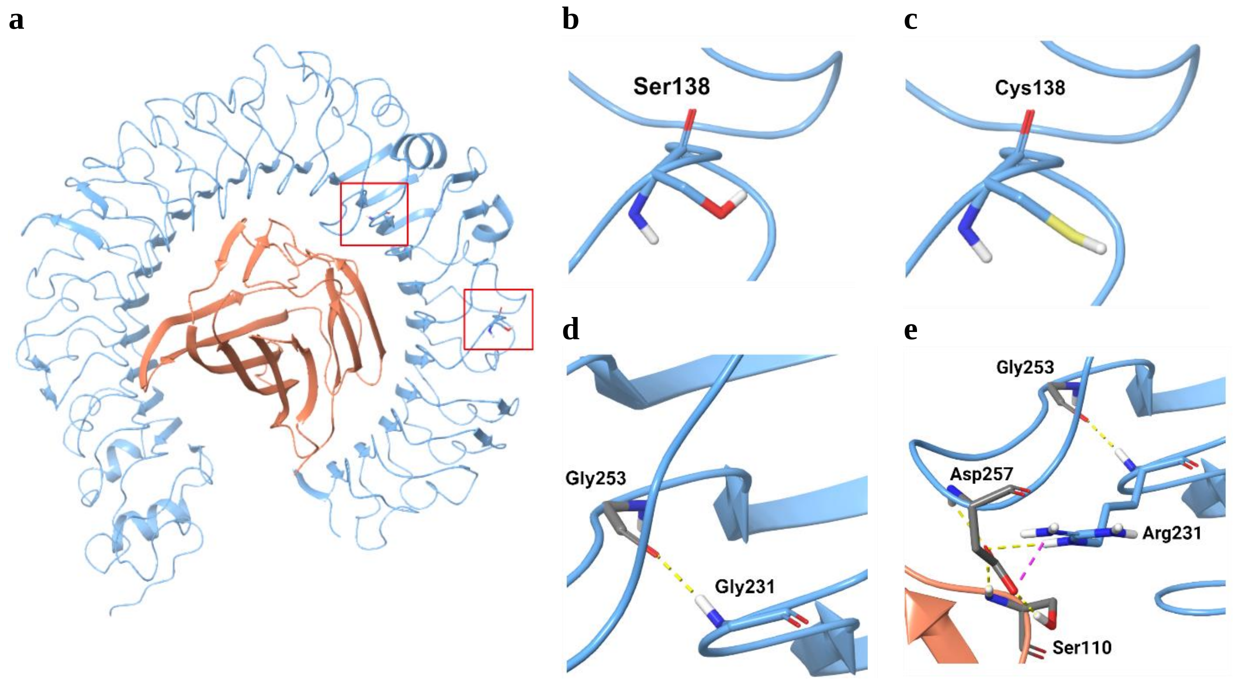


**Supplementary Figure S24. Modeled 3D structure of CD180 bound to MD1.** The CD180 and MD1 proteins are shown in blue and orange cartoon forms, respectively and amino acids are represented with stick representations. The red-boxed regions shown in (a) are magnified in successive images. (a) 3D crystal structure of CD180-MD1 complex; (b) Wild type Ser138; (c) Mutant Cys138; (d) Wild type Gly231; (e) Mutant Arg231.

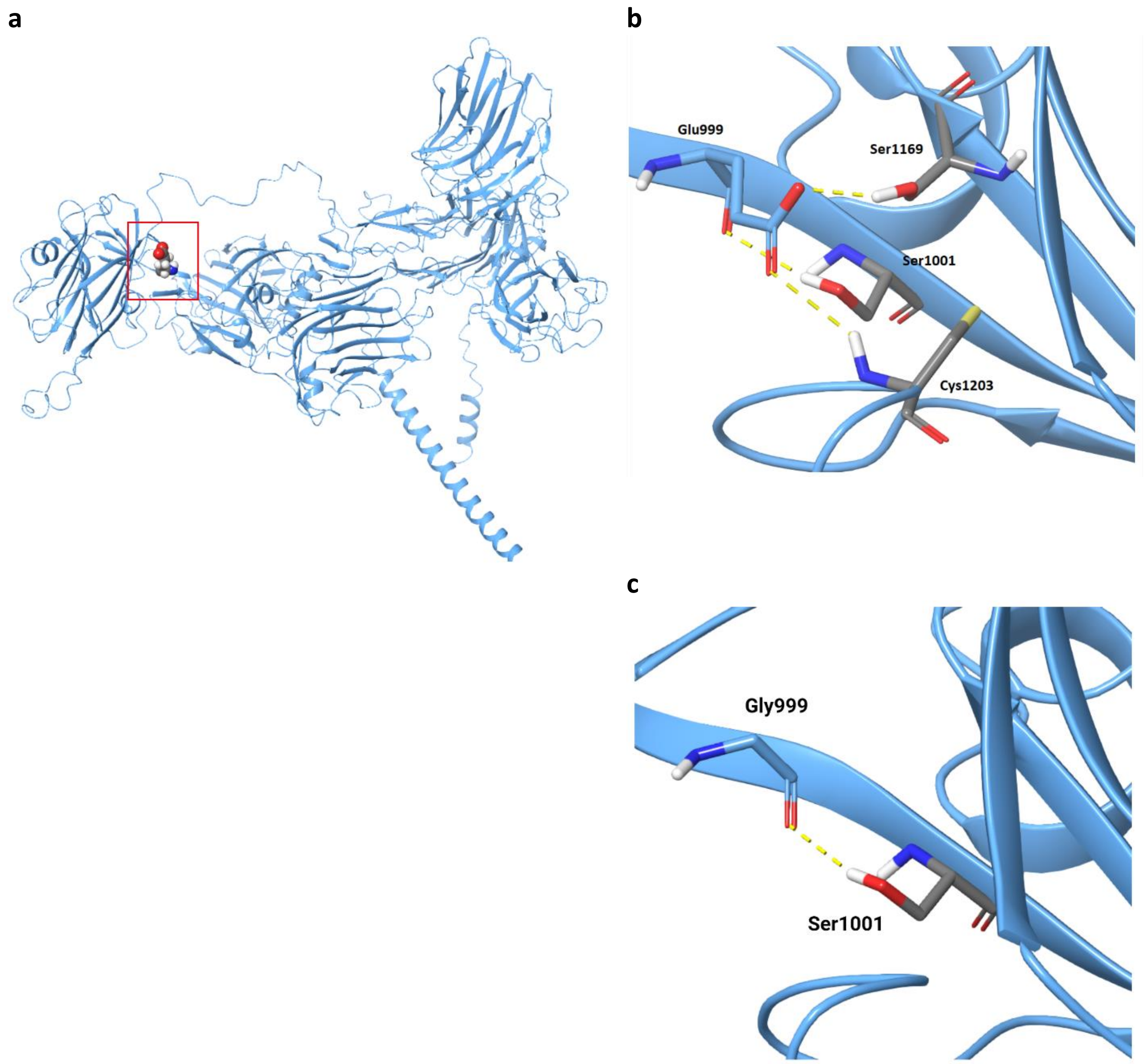


**Supplementary Figure S25. Modeled 3D structure of CNTNAP3.** Amino acids are represented with sticks. The red-boxed region in (a) is enlarged in the successive images (b and c). (a) Modeled structure of CNTNAP3; (b) Wild type Glu999; (c) Mutant Gly999.

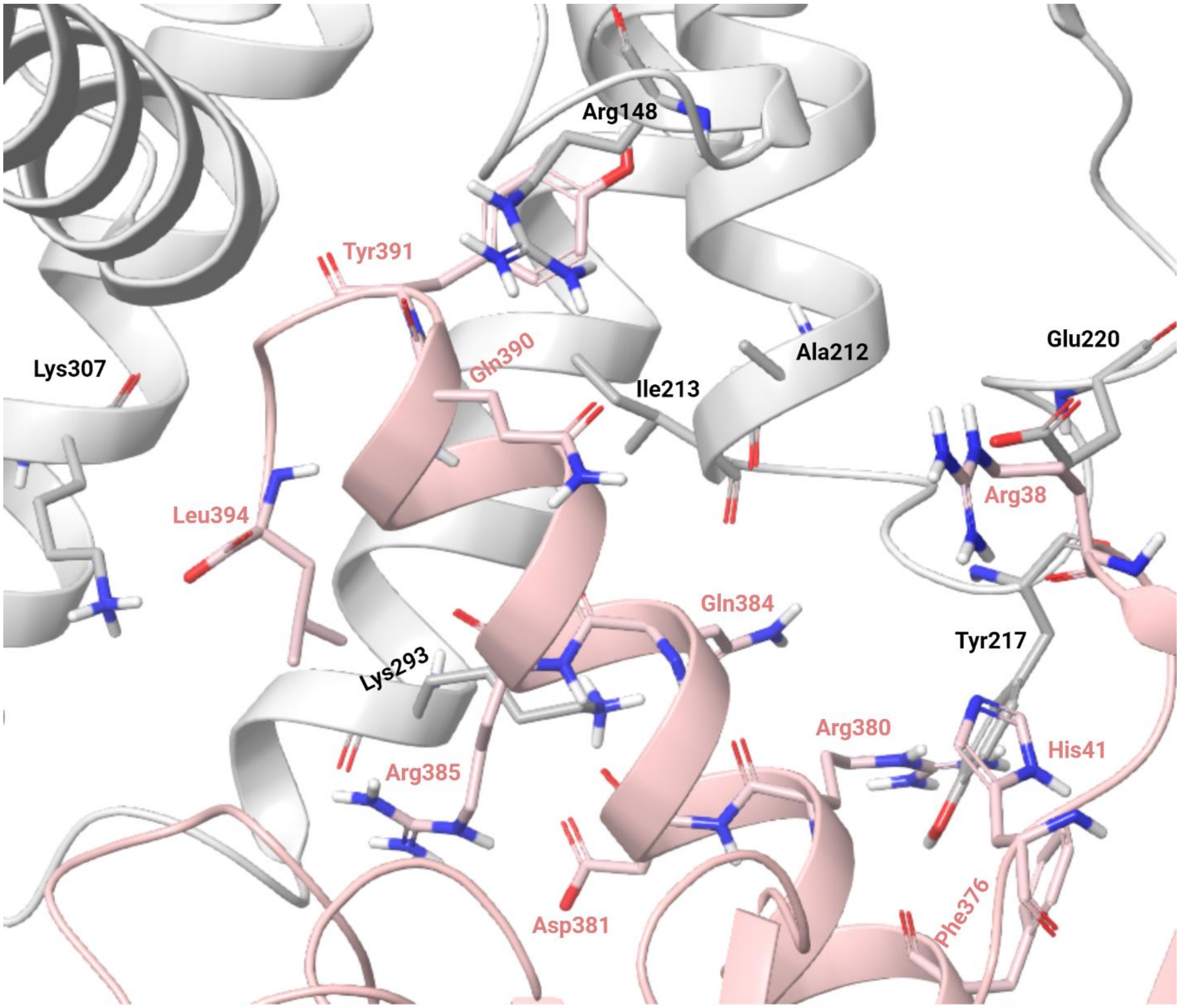


**Supplementary Figure S26. A close-up view of the binding interface of CRHR2 and Gα protein.** The interacting residues of CRHR2 and Gα are shown in grey and pink stick representations, respectively.

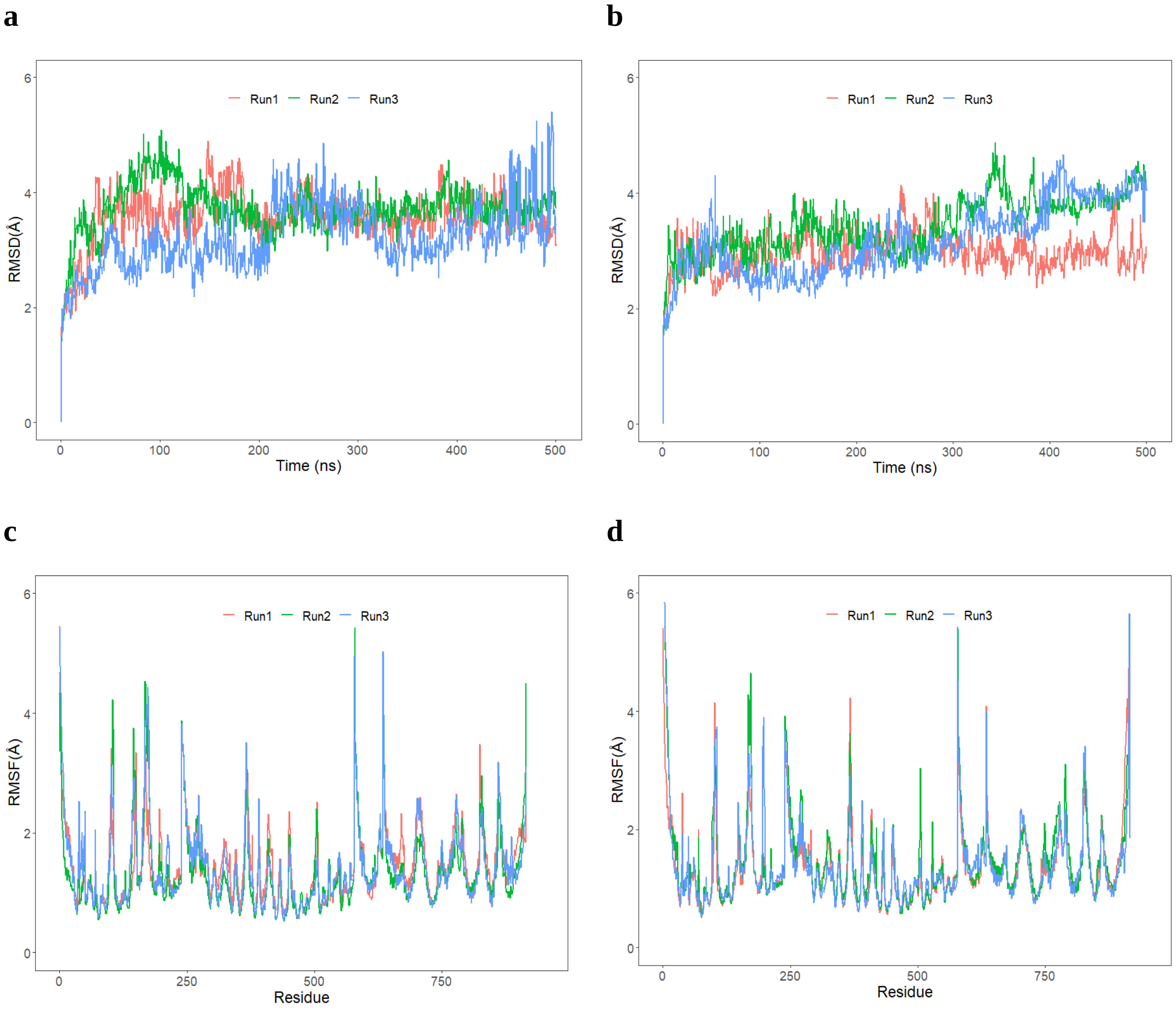


**Supplementary Figure S27. RMSD and RMSF plots of triplicate 500 ns simulations of CRHR2.** Data from the three runs are plotted with red, blue and green lines. a) RMSD of protein Cα atoms from the wild type CRHR2 simulations. b) RMSD of protein Cα atoms from the mutant CRHR2 simulations. c) RMSF of protein Cα atoms from the wild type CRHR2 simulations. d) RMSF of protein Cα atoms from the mutant CRHR2 simulations.

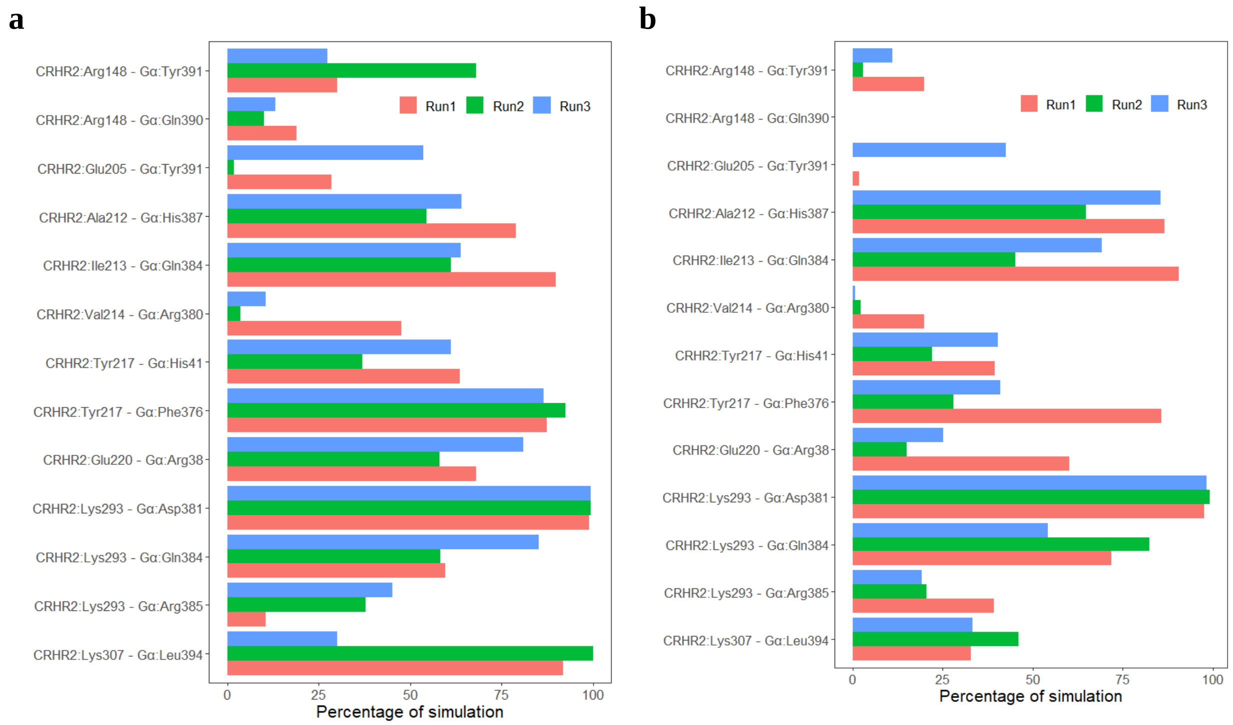


**Supplementary Figure S28. The percentage of simulation time during which intermolecular contacts were retained between CRHR2 and Gα protein**. (a) Intermolecular contacts from three independent runs of wild type CRHR2–Gα structures. (b) Intermolecular contacts from three independent runs of mutant CRHR2–Gα structures.

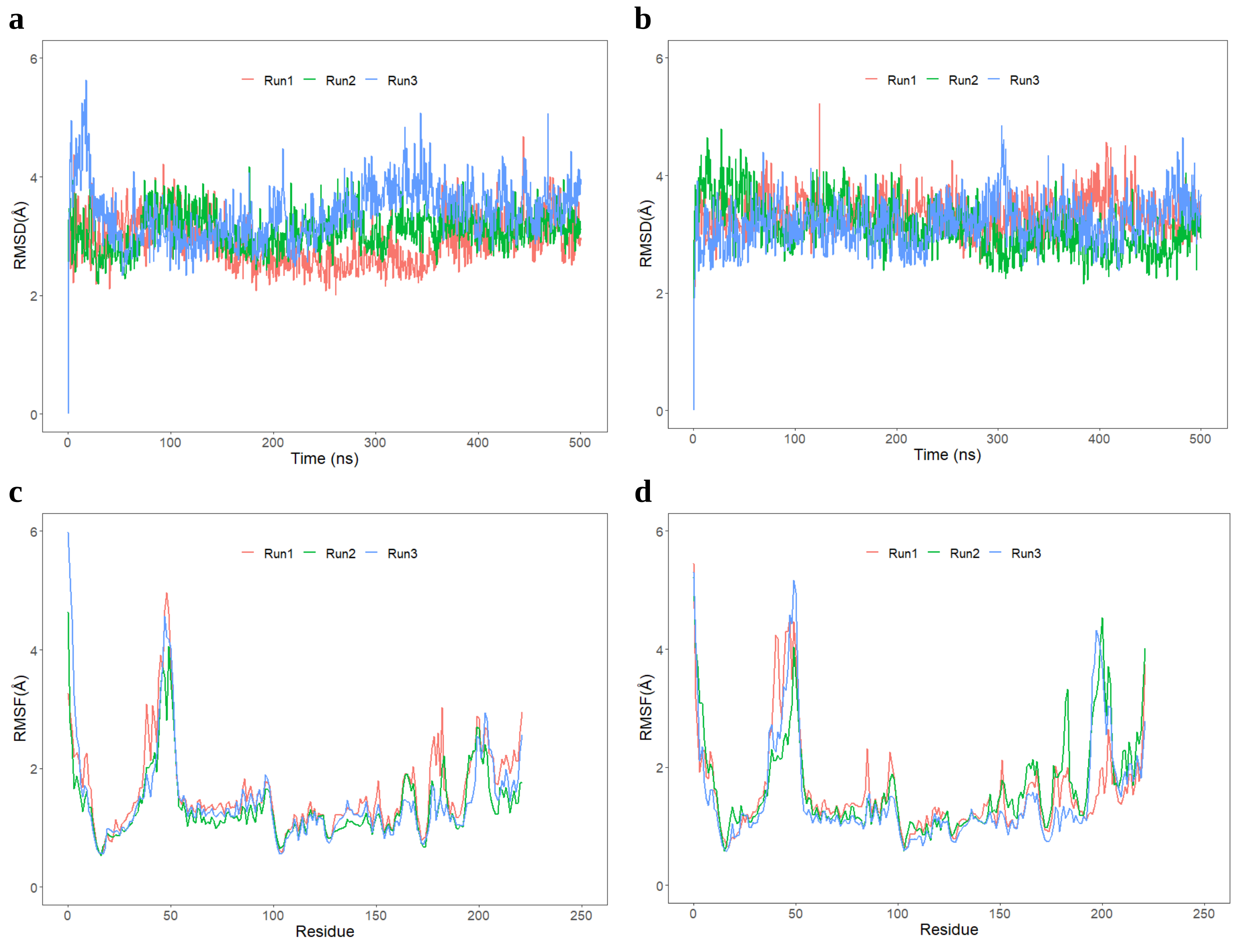


**Supplementary Figure S29. RMSD and RMSF plots of triplicate 500 ns simulations of PSPH.** Data from the three runs are plotted with red, blue and green lines. a) RMSD of protein Cα atoms from the wild type PSPH simulations. b) RMSD of protein Cα atoms from the mutant PSPH simulations. c) RMSF of protein Cα atoms from the wild type PSPH simulations. d) RMSF of protein Cα atoms from the mutant PSPH simulations.

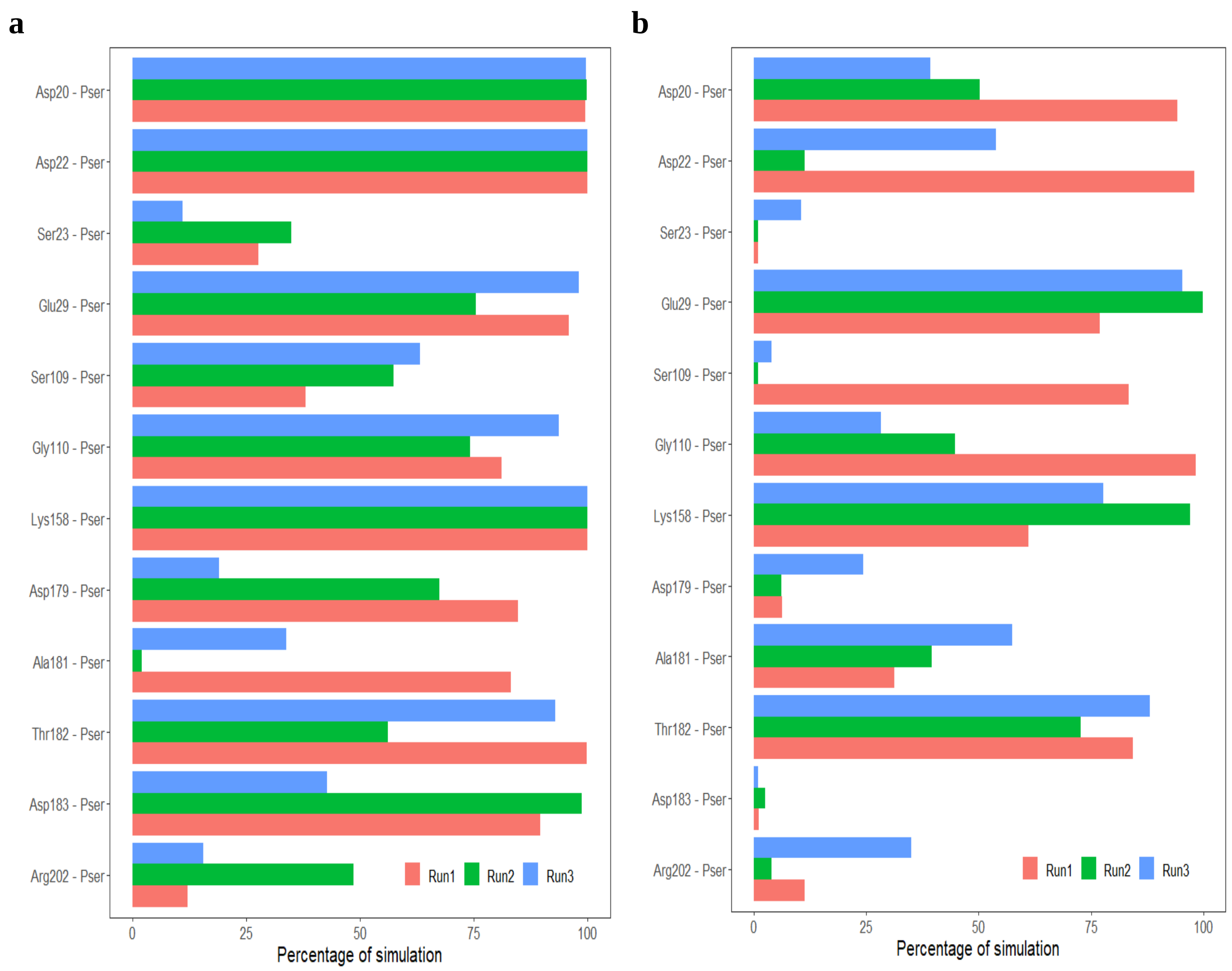


**Supplementary Figure S30. The percentage of simulation time during which intermolecular contacts were retained between PSPH and phosphoserine (Pser).** (a) Intermolecular contacts from three independent runs of wild type PSPH–Pser complex. (b) Intermolecular contacts from three independent runs of mutant PSPH–Pser complex.

**Supplementary Table S1. Data on study cohorts**

| Characteristic | Common variant association study cohort | Rare variant association study cohort |
|---|---|---|
| Number of subjects | 742 | 632 |
| Female (%) | 53.8 | 55.1 |
| Baseline age, mean ± SD (range) | 75.0 ± 6.6 (61.9 to 94.3) | 74.7 ± 6.4 (61.9 to 94.3) |
| Follow-up time, years (mean ± SD) | 6.54 ± 3.4 | 7.0 ± 3.2 |

**Supplementary Table S2. 18 putative risk rare coding variants for episodic memory decline implicated by single rare variant association analysis**

| Variant | Gene | Annotation | Carriers | CADD | Coefficient | P | FDR |
|---|---|---|---|---|---|---|---|
| 17:4049847:A:G (L1959P) | *ZZEF1* | missense | 6 | 22.5 | -0.069 | 1.12E-06 | 0.0016 |
| 1:2592689:A:G (I678T) | *MMEL1* | missense | 5 | 25.2 | -0.069 | 6.61E-06 | 0.0038 |
| 2:140902978:C:G (K1236N) | *LRP1B* | missense | 6 | 23.6 | -0.062 | 7.89E-06 | 0.0038 |
| 9:39088647:T:C (E999G) | *CNTNAP3* | splice, missense | 11 | 26.1 | -0.044 | 2.32E-05 | 0.0092 |
| 6:116792606:C:T (G773S) | *GPRC6A* | missense | 11 | 25.5 | -0.043 | 3.58E-05 | 0.014 |
| 6:116792609:T:C (K772E) | *GPRC6A* | missense | 11 | 25.8 | -0.043 | 3.58E-05 | 0.014 |
| 15:83892716:T:C (V432A) | *ADAMTSL3* | missense | 5 | 23.2 | -0.061 | 7.99E-05 | 0.026 |
| 1:48235797:A:G (S429G) | *SLC5A9* | missense | 8 | 26.8 | -0.048 | 1.0E-04 | 0.026 |
| 16:88628759:C:T (A848V) | *ZC3H18* | splice, missense | 5 | 24.8 | -0.059 | 1.16E-04 | 0.026 |
| 5:157343406:A:T (L44H) | *FNDC9* | missense | 12 | 26.2 | -0.038 | 1.18E-04 | 0.026 |
| 7:56015138:G:A (T152I) | *PSPH* | missense | 8 | 27.9 | -0.047 | 1.31E-04 | 0.029 |
| 12:132572582:C:T (P515L) | *FBRSL1* | missense | 10 | 23.7 | -0.041 | 1.34E-04 | 0.029 |
| 12:132581511:T:C (L679P) | *FBRSL1* | missense | 10 | 24.6 | -0.041 | 1.34E-04 | 0.029 |
| 17:63941396:G:A (P1629L) | *SCN4A* | missense | 8 | 25.4 | -0.047 | 1.35E-04 | 0.029 |
| 17:42039863:G:A (R624W) | *C17orf113* | missense | 11 | 26.1 | -0.04 | 1.52E-04 | 0.033 |
| 17:42544215:C:A (R737S) | *NAGLU* | missense | 12 | 20.9 | -0.038 | 1.61E-04 | 0.034 |
| 12:109904025:G:A (E93K) | *TCHP* | missense | 5 | 23.2 | -0.057 | 2.13E-04 | 0.043 |
| 17:42300824:G:A (E315K) | *STAT5A* | missense | 5 | 22.6 | -0.057 | 2.33E-04 | 0.046 |

**Supplementary Table S3. Rare variants included in gene-based association test for 6 significant genes.**

| Variant | Gene | Annotation | Carriers | CADD | Coefficient | *P* |
|---|---|---|---|---|---|---|
| 21:33750170:C:A | *ITSN1* | missense | 1 | 22.3 | 0.037 | 0.276 |
| **21:33750235:G:A** | ***ITSN1*** | **missense** | **3** | **23.7** | **-0.076** | **3.55E-08** |
| 21:33750307:G:C | *ITSN1* | missense | 1 | 25.4 | -0.018 | 0.599 |
| **21:33772240:G:T** | ***ITSN1*** | **stop_gained** | **1** | **38** | **-0.098** | **0.004** |
| 21:33836464:G:A | *ITSN1* | missense | 1 | 29.4 | 0.018 | 0.607 |
| 5:67184151:C:T | *CD180* | missense | 1 | 23.2 | 0.027 | 0.427 |
| **5:67184152:C:T** | ***CD180*** | **missense** | **2** | **23.2** | **-0.111** | **4.68E-06** |
| **5:67184430:G:C** | ***CD180*** | **missense** | **2** | **24.5** | **-0.067** | **0.005** |
| 5:67185998:T:C | *CD180* | missense | 1 | 24.4 | -0.007 | 0.848 |
| 7:30662733:C:T | *CRHR2* | missense | 1 | 24.1 | -0.003 | 0.933 |
| **7:30665171:G:A** | ***CRHR2*** | **missense** | **4** | **27.7** | **-0.077** | **5.81E-06** |
| 7:30665594:A:T | *CRHR2* | missense | 1 | 23.1 | -0.054 | 0.116 |
| 17:4013589:C:A | *ZZEF1* | missense | 1 | 23.1 | 0.058 | 0.093 |
| **17:4049847:A:G** | ***ZZEF1*** | **missense** | **6** | **22.5** | **-0.069** | **1.12E-06** |
| 17:4050793:G:A | *ZZEF1* | missense | 4 | 23.7 | -0.003 | 0.870 |
| 17:4050814:C:T | *ZZEF1* | missense | 1 | 25.9 | 0.042 | 0.224 |
| 17:4072622:G:A | *ZZEF1* | missense | 1 | 23.9 | -0.023 | 0.506 |
| 17:4095975:C:T | *ZZEF1* | missense | 2 | 28.4 | 0.037 | 0.128 |
| 17:4114401:T:C | *ZZEF1* | missense | 3 | 24.4 | 0.015 | 0.450 |
| 17:4142577:G:C | *ZZEF1* | missense | 1 | 31 | 0.024 | 0.482 |
| 17:4142815:C:G | *ZZEF1* | missense | 1 | 24.6 | -0.041 | 0.233 |
| 15:41835004:G:A | *JMJD7* | missense | 5 | 23.9 | -0.021 | 0.170 |
| **15:41835094:G:A** | ***JMJD7*** | **missense** | **1** | **24.3** | **-0.069** | **0.044** |
| **15:41836175:A:G** | ***JMJD7*** | **missense** | **2** | **29.7** | **-0.083** | **0.001** |
| **15:41836243:G:C** | ***JMJD7*** | **splice, missense** | **2** | **33** | **-0.075** | **0.002** |
| 1:2591979:C:T | *MMEL1* | missense | 1 | 23.7 | 0.010 | 0.761 |
| **1:2592689:A:G** | ***MMEL1*** | **missense** | **5** | **25.2** | **-0.069** | **6.62E-06** |
| 1:2593904:A:AG | *MMEL1* | frameshift | 1 | 34 | -0.002 | 0.957 |

| | | | | | | |
|---|---|---|---|---|---|---|
| 1:2603925:G:A | *MMEL1* | missense | 1 | 22.5 | -0.015 | 0.672 |

Note:

1. Variants in bold are putative risk variants underlying the significant rare-variant gene-based association.
2. *P* denotes single variant association *P* value.
3. The putative risk variants in *MMEL1* and *ZZEF1* were already identified by the single-rare variant association analysis.

**Supplementary Table S4. Protein stability analysis of rare coding variants.**

| Gene | Variant | Stability* (kcal/mol) |
|---|---|---|
| *ZZEF1* | ZZEF1:p.L1959P | 36.79 |
| *MMEL1* | MMEL1:p.I678T | 16.9 |
| *CNTNAP3* | CNTNAP3:p.E999G | 9.87 |
| *GPRC6A* | GPRC6A:p.K772E | -9.81 |
| *GPRC6A* | GPRC6A:p.G773S | 1.34 |
| *ADAMTSL3* | ADAMTSL3:p.V432A | 4.26 |
| *SLC5A9* | SLC5A9:p.S429G | 7.74 |
| *FNDC9* | FNDC9:p.L44H | 4.69 |
| *PSPH* | PSPH:p.T152I | 3.87 |
| *SCN4A* | SCN4A:p.P1629L | 2.65 |
| *C17orf113* | C17orf113:p.R624W | 31.31 |
| *NAGLU* | NAGLU:p.R737S | 22.52 |
| *STAT5A* | STAT5A:p.E315K | 2.61 |
| *JMJD7* | JMJD7:p.V115M | -3.25 |
| *JMJD7* | JMJD7:p.Y186C | 23.11 |
| *JMJD7* | JMJD7:p.E209Q | -2.42 |
| *CD180* | CD180:p.G231R | 23.12 |
| *CD180* | CD180:p.S138C | -3.16 |
| *CRHR2* | CRHR2:p.R148W | 31.94 |
| *TCHP* | TCHP:p.E93K | 15.27 |
| *LRP1B* | LRP1B:p.K1236N | 1 |
| *ITSN1* | ITSN1:p.V147I | -0.23 |
| *FBRSL1* | FBRSL1:p.P515L | -3.14 |
| *FBRSL1* | FBRSL1:p.L679P | 31.2 |
| *ZC3H18* | ZC3H18:p.A848V | 3.49 |

Note:

1. *A positive value indicates that the mutant protein is less stable than the wild-type protein; a negative value indicates that the mutant protein is more stable that the wild-type protein.
2. Only missense variants were analyzed.

### Supplementary Table S5. List of studied variants at protein level and their impact on respective protein's intermolecular interactions.

| Gene | Variant (interfacial variants in bold) | Type | PDB[1] | Template | Identity (%) | AlphaFold ID | PTM site[2] | Residues forming hydrogen bonds with wild type | Residues forming hydrogen bonds with mutant |
|---|---|---|---|---|---|---|---|---|---|
| *ZZEF1* | 17:4049847:A>G; p.L1959P | Missense | N/A | 7Z8B | 41.2 | | No | Leu1811 | Leu1811 |
| *MMEL1* | 1:2592689:A>G; p.I678T | Missense | N/A | | | AF-Q495T6-F1 | No | Gly682 | Leu674, Gly682 |
| *LRP1B* | 2:140902978:C>G; p.K1236N | Missense | N/A | 1N7D | 49 | | No | Glu1229, Cys1237 | Glu1229 |
| *CNTNAP3* | 9:39088647:T>C; p.E999G | Missense | N/A | | | AF-Q9BZ76-F1 | No | Ser1001, Ser1169, Cys1203 | Ser1001 |
| *GPRC6A* | 6:116792609:T>C; p.K772E | Missense | N/A | | | AF-Q5T6X5-F1 | No | Ile768 | Ile768, Tyr775 |
| *GPRC6A* | 6:116792606:C>T; p.G773S | Missense | N/A | | | AF-Q5T6X5-F1 | No | Phe769, Glu776 | Phe769, Glu776 |
| *ADAMTSL3* | 15:83892716:T>C; p.V432A | Missense | N/A | | | AF-P82987-F1 | No | Asn475 | Asn475 |
| *SLC5A9* | 1:48235797:A>G; p.S429G | Missense | N/A | | | AF-Q2M3M2-F1 | No | Gln303, Ser400, Leu408 | Ser400, Leu408 |
| *FNDC9* | 5:157343406:A>T; p.L44H | Missense | N/A | | | AF-Q8TBE3-F1 | No | Trp36 | Trp36 |
| *PSPH* | **7:56015138:G>A; p.T152I** | Missense | 1L8L | | | | No | Phe58, Ser109 | Phe58 |
| *SCN4A* | 17:63941396:G>A; p.P1629L | Missense | 6MBA | | | | No | Nil | Nil |
| *C17orf113* | 17:42039863:G>A; p.R624W | Missense | N/A | | | AF-A0A1B0GUU1-F1 | No | Thr209, Ser239, Ser620, Gly627, Gln628 | Ser620, Gly627, Gln628, Arg635 |
| *NAGLU* | 17:42544215:C>A; p.R737S | Missense | 4XWH | | | | No | Tyr734, Gly741 | Gly741, Ser742 |
| *TCHP* | 12:109904025:G>A; p.E93K | Missense | N/A | | | AF-Q9BT92-F1 | No | Gln89, Leu97 | |
| *STAT5A* | 17:42300824:G>A; p.E315K | Missense | 7TVA | | | | No | Glu311, Thr319 | Glu311, Thr319 |
| *CD180* | 5:67184430:G>C; p.S138C | Missense | 3B2D | | | | No | | |
| *CD180* | **5:67184152:C>T; p.G231R** | Missense | 3B2D | | | | No | CD180:Gly253 | CD180:Gly253, Asp257 MD:Ser110 |
| *CRHR2* | **7:30665171:G>A; p.R175W** | Missense | 6PB1 | | | | No | Gα:Gln390; CRHR2:His152, CRHR2:Glu365 | CRHR2:His152, CRHR2:Glu365 |
| *ITSN1* | 21:33750235:G>A; p.V147I | Missense | - | Reliable template not available | | | No | | |
| *JMJD7* | 15:41835094:G>A; p.V115M | Missense | 5NFN | | | | No | Phe111, Gly118 | Phe111, Gly118 |
| *JMJD7* | 15:41836175:A>G; p.Y186C | Missense | 5NFN | | | | No | Phe195, Leu269 | Leu269 |
| *JMJD7* | 15:41836243:G:C; p.E209Q | Missense | 5NFN | | | | No | Trp237, Tyr250 | Trp237, Tyr250 |

Note:
1. PDB, structure from Protein Data Bank
2. PTM, Posttranscriptional modification
